# Abnormal higher-order network interactions in Parkinson’s disease visual hallucinations

**DOI:** 10.1101/2023.04.11.23288391

**Authors:** Joshua B. Tan, Eli J. Müller, Isabella F. Orlando, Natasha L. Taylor, Daniel S. Margulies, Jennifer Szeto, Simon J.G. Lewis, James M. Shine, Claire O’Callaghan

**Author notes:** **Correspondence to:** Claire O’Callaghan Brain and Mind Centre, 94 Mallett Street, Camperdown NSW 2050, Australia.

## Abstract

Visual hallucinations in Parkinson’s disease can be viewed from a systems-level perspective, whereby abnormal communication between brain networks responsible for perception predisposes a person to hallucinate. To this end, abnormal functional interactions between higher-order and primary sensory networks have been implicated in the pathophysiology of visual hallucinations in Parkinson’s disease, however the precise signatures remain to be determined. Dimensionality reduction techniques offer a novel means for simplifying the interpretation of multidimensional brain imaging data, identifying hierarchical patterns in the data that are driven by both within- and between-functional network changes. Here, we applied two complementary non-linear dimensionality reduction techniques – diffusion-map embedding and t-distributed Stochastic Neighbour Embedding (t-SNE) – to resting state fMRI data, in order to characterise the altered functional hierarchy associated with susceptibility to visual hallucinations. Our study involved 77 people with Parkinson’s disease (31 with hallucinations; 46 without hallucinations) and 19 age-matched healthy controls. In patients with visual hallucinations, we found compression of the unimodal-heteromodal gradient consistent with increased functional integration between sensory and higher order networks. This was mirrored in a traditional functional connectivity analysis, which showed increased connectivity between the visual and default-mode networks in the hallucinating group. Together, these results suggest a route by which higher-order regions may have excessive influence over earlier sensory processes, as proposed by theoretical models of hallucinations across disorders. By contrast, the t-SNE analysis identified distinct alterations in prefrontal regions that were not apparent in the functional connectivity analysis, suggesting complex reconfigurations in functional brain network architecture as a function of the disease process. Together, the results confirm abnormal brain organisation associated with the hallucinating phenotype in Parkinson’s disease, and highlight the utility of applying convergent dimensionality reduction techniques to investigate complex clinical symptoms. In addition, the patterns we describe in Parkinson’s disease converge with those seen in other conditions, suggesting that reduced hierarchical differentiation across sensory-perceptual systems may be a common transdiagnostic vulnerability in neuropsychiatric disorders with perceptual disturbances.

## Introduction

Veridical perception requires the ability to interact with and process a continuous stream of sensory information. These interactions rely on associations developed over time, whereby perceptual interpretations are informed by matching sensory inputs with statistics learned about features of the external environment.^1, 2^ In this framework, hallucinations are proposed to occur due to an imbalance between higher-order (“top-down”) vs. sensory (“bottom-up”) processes.^3–6^ More concretely, hallucinations have been proposed to arise due to disruptions across networks involved in higher-order perceptual processing (i.e., attentional networks, the default mode network) and primary sensory networks.^4, 7^ In clinical populations with visual hallucinations, including Parkinson’s disease and Lewy body dementia, abnormal interactions within and between these networks have been consistently observed.^6–9^ In this way, a systems-level perspective that focuses on dysfunctional patterns of communication between brain networks can provide insight into the neural signatures of visual hallucinations.

Tracking neural activity during hallucinatory episodes is notoriously difficult, but trait-level signatures of the tendency to hallucinate can be explored using structural imaging or resting state fMRI.^10–12^ These approaches identify patterns of abnormal brain structure, activity or connectivity associated with the hallucinating phenotype, which are presumably implicated in hallucinatory events. For example, the network abnormalities observed during fMRI of hallucination-like events in Parkinson’s disease overlaps with trait-level network abnormalities observed in the resting state.^8, 13, 14^ However, one challenge is that resting state fMRI patterns are inherently high-dimensional – i.e., the data has an extensive and unwieldy number of features – which poses issues for interpretability and reproducibility. A tractable way to handle this complexity is to apply dimensionality reduction techniques, which are algorithms that extract latent components from high-dimensional data while preserving relationships of the original data^15^ and discarding more idiosyncratic features.^15, 16^ This approach offers a means of summarising feature-rich data into components that can then be more meaningfully related to symptoms and behaviour.

One popular method for reducing dimensionality is diffusion map embedding – a non-linear dimensionality reduction technique, which projects high-dimensional data into an n-dimensional gradient space where n ≤ the number of data points.^17, 18^ In the case of resting-state fMRI, the resulting “map” of brain activity represents the global connectivity structure as a distribution of cortical nodes: nodes that share stronger connections are grouped closer together, whereas nodes that do not share connections are grouped further apart.^19, 20^ Diffusion map embedding has been used to demonstrate a key organisational principle in healthy human brains that links “bottom-up”, sensory (unimodal) regions with “top-down”, higher-order (heteromodal) cortical areas along a primary gradient.^21, 22^

The unimodal-heteromodal gradient is widely replicated across studies and populations,^21, 22^ and is sensitive to age-related changes^23^ and clinical conditions, including autism^24^ and schizophrenia.^25^ Specifically, a reduced separation (i.e., a compression) along the gradient between sensory and higher-order regions is seen in neuropsychiatric patient groups relative to controls.^24, 25^ In Parkinson’s disease, this unimodal-heteromodal gradient has been shown to be compressed in patients with visual dysfunction.^26^ It follows that increased functional integration between previously well-separated sensory and higher-order regions (i.e., primary sensory and default mode regions) may reflect abnormal interactions between such regions. These changes could potentially disturb perceptual processes, allowing for an increased influence from higher-order regions over lower-level sensory processes – increasing the vulnerability to hallucinate. Taken together, changes in the hierarchical organisation of the unimodal-heteromodal gradient may serve as a transdiagnostic feature across neuropsychiatric disorders. In turn, alterations in this unimodal-heteromodal gradient organisation may be an underlying feature that helps explain the network disruptions observed in Parkinson’s disease patients prone to visual hallucinations.

A pitfall of dimensionality reduction techniques is that they require simplifying assumptions, which can obscure interpretation of the underlying functional neuroanatomy. One solution is to use multiple approaches, each with their own strengths and weaknesses, to converge on a plausible interpretation of the data. In contrast to diffusion map embedding, t-distributed Stochastic Neighbour Embedding (t-SNE)^27, 28^ computes a similarity score between all data points in a high-dimensional space, and then maps these similarities into a lower (typically 2-3) dimensional space. In this way, t-SNE allows for visual interrogation of network organisation,^29, 30^ while conserving the relationships between data points^29, 31^, albeit in a different way than diffusion-map embedding that is potentially more sensitive to non-linear reconfigurations in network architecture (i.e., t-SNE captures both local and global features, whereas diffusion map embedding only focuses on local features). Combining diffusion-map embedding and t-SNE thus has the potential to expose the higher-order organisation of resting-state networks, and offer unique insights into the changes in network topology brought on by neurodegenerative disease processes.

Here, we combine diffusion map embedding and t-SNE to determine the low-dimensional signature of the tendency to hallucinate in individuals with Parkinson’s disease. To do so, we analysed resting-state fMRI data from Parkinson’s disease patients with visual hallucinations compared to those without, along with age-matched healthy controls. We hypothesised that patients with visual hallucinations would show compression in their unimodal-heteromodal gradient, and the extent of gradient compression would be associated with cognitive decline. We also predicted that compression in the gradient would be complemented by a decreased distance between subsets of the whole-brain network as detected through t-SNE analysis.

## Materials and methods

### Case selection

A total of 96 individuals were recruited from the Parkinson’s disease Research Clinic at the Brain and Mind Centre, University of Sydney, Australia, including 19 healthy controls and 77 people diagnosed with idiopathic Parkinson’s disease. All Parkinson’s disease patients satisfied the United Kingdom Parkinson’s Disease Society Brain Bank criteria and did not meet criteria for dementia.^32^ Parkinson’s disease symptoms were assessed with the Movement Disorder Society-Sponsored Revision of the Unified Parkinson’s Disease Rating Scale (MDS-UPDRS).^33^ Patients with visual hallucinations were identified from a positive response to question two of the MDS-UPDRS (i.e., “Over the past week have you seen, heard, smelled, or felt things that were not really there? If yes, examiner asks the patient or caregiver to elaborate and probes for information”). Individuals scoring ≥1 on this item, with a subsequent description consistent with visual hallucinatory phenomena, were included in the hallucinating group. Thirty-one patients were identified as experiencing visual hallucinations and 46 did not experience visual hallucinations. All patients were tested on their regular dopamine medications and a dopaminergic dose equivalent (DDE) score was calculated (mg dopamine/per day).^34^ Psychiatric symptoms were screened using the Scales for Outcomes in Parkinson’s Disease - Psychiatric Complications (SCOPA-PC),^35^ and a subset of 47 patients (20 with hallucinations, 27 without) underwent the Psychosis and Hallucinations Questionnaire (PsycH-Q).^36^

### Neuropsychological and behavioural assessments

Global cognition was assessed via the Mini-Mental State Examination (MMSE)^37^ and Montreal Cognitive Assessment (MoCA).^38^ The Trail Making Test parts A and B (TMT-A, TMT-B) measured psychomotor speed and attentional set shifting capacity (TMT-B minus TMT-A).^39^ Working memory maintenance and manipulation was assessed using the Digit Span Test,^40^ consisting of two parts: digit spans forwards and backwards, which were summed to create a digit span total score (Digit Span Total; DST). Memory was assessed via the percentage of a short story correctly recalled after 30 minutes (Logical Memory retention: LM Retention).^40^

### Statistical analysis

Demographic analyses were performed in R version 4.2.1.^41^ For missing scores in the cognitive dataset, data imputation was conducted using Multivariate Imputation via Chained Equations (MICE) from the ‘mice’ package.^42^ As the missing values all belonged to quantitative variables, predictive mean matching was used. For all analyses, the group comparisons focused on the Parkinson’s disease group as a whole versus controls (PD vs. controls) and within the Parkinson’s group, hallucinators versus non-hallucinators (PD+VH vs. PD–VH). Group comparisons were conducted through non-parametric permutation testing (5000 permutations).

### MRI acquisition

All 96 individuals underwent magnetic resonance imaging (MRI) on a 3-Tesla MRI scanner (GE medical systems), generating T1-weighted structural images and resting-state blood-oxygenation level dependent (BOLD) functional scans (rsfMRI). Sagittal 3D T1w were acquired using a 256 × 256 matrix, 200 slices, slice thickness of 1 mm, echo time/repetition time = 2.7/7.2 ms. Functional images were acquired with repetition time = 3 s, echo time = 36 ms, flip angle = 90°, 32 axial slices covering the whole brain, field of view = 220 mm, slice thickness of 3 mm, raw voxel size = 3.9 mm × 3.9 mm × 4 mm, and 140 repetition times (scanning duration of 7 min). Individuals were instructed to lie awake with their eyes open.

### MRI preprocessing

Scans were converted into the Brain Imaging Data Structure^43^ format using the dicm2nii^44^ and dicm2niix^45^ toolboxes. Preprocessing was completed using *fMRIPrep* 20.2.3,^46^ a standard pipeline that incorporates toolboxes from the gold-standard preprocessing software in the field. *fMRIPrep* involves the basic preprocessing steps (coregistration, normalisation, unwarping, noise component extraction, segmentation, skullstripping etc.) and produces a report for quality checking at each step. See Supplementary Material for a full description of each step.

### Denoising

The confounds timeseries data extracted from *fMRIprep* were passed through *fmridenoise*^47^ specifying eight physiological signals to be regressed (mean physiological signals from white matter and cerebrospinal fluid, and their quadratic terms^48^), with high-pass and low-pass band filters set at 0.01 and 0.1 respectively.

### Gradient connectivity analysis

Mean BOLD signal time-series data were extracted from the rsfMRI data for 400 cortical regions from the Schaefer atlas^49^ and z-scored, using MATLAB scripts adapted from the fieldtrip toolbox.^50^ A functional connectivity matrix was calculated for each individual using Pearson correlation values, producing a 400 × 400 matrix that represented cortical-cortical functional connectivity. These 400 cortical regions were assigned to 7 resting state networks,^51^ allowing for comparisons between the cortical regions and large-scale cortical networks.^49^

Gradient analysis was performed using the *Brainspace* toolbox and custom MATLAB scripts.^17^ First, a population average connectivity matrix was calculated using the extracted timeseries data from all the individual 400 × 400 connectivity matrices. The average matrix was thresholded, with the top 10% of measurements per row retained and all remaining measurements zeroed. An affinity matrix was then computed using the normalised angle method – this reflected the similarity of connectivity profiles between each pair of regions. Then, diffusion map embedding was used to simplify the high-dimensional nature of the data into lower dimensions, allowing for components to be generated in descending order from highest to lowest variance explained. The density of sampling points was controlled through the parameter α = 0.5, following recommendations from previous studies, retaining global relations between the data points in the embedded space.^17, 19^

Gradient components were calculated for each individual using the same parameters as the group-level average gradient. Individual gradients were then aligned to the group-level gradient using Procrustes alignment,^19^ allowing for more accurate comparisons across individuals. The first and second gradients which explained most variance in the data (14% and 12%, respectively) were extracted and compared against the presumed network hierarchy as a comparison against the principal gradient described by Margulies and colleagues.^22, 26, 52^

### Comparison of unimodal-heteromodal gradient with behavioural data

To determine whether changes in the gradient score were associated with cognitive performance, for each individual we calculated the average gradient score for each network in the Yeo 7-network atlas.^51^ We focused on networks significantly different between the three groups in our cohort (i.e., visual, ventral attentional, and frontoparietal control) and correlated those network gradient scores with clinical scores that were significantly different between the patient groups (i.e., TMT-B, BDI, and HADS).

### t-Stochastic Neighbour Embedding (t-SNE) analysis

The t-SNE algorithm in MATLAB^53^ was used to construct 3-dimensional embeddings of each individual functional connectivity matrix. Before running the data through the t-SNE algorithm, the data underwent a PCA initialisation step in which the top 3 components were selected.^54^ This resulted in a 400 × 3 matrix where each of the 400 cortical regions was described by x-y-z coordinates. The algorithm was run for 1,000 iterations using the Barnes-Hut algorithm which performs an approximate optimisation. The distance metric was set to “Euclidean”; perplexity = 90; learning rate = 500; exaggeration = 50 for the first 99 optimisations to facilitate cluster formation.^31^ For specific details regarding the choice of parameter values refer to the Supplementary Materials. A t-SNE map was generated for each individual using the parameters specified above. For each individual t-SNE map, the Euclidean distance was calculated between each pair of cortical regions generating a “distance map” that described how far each region was from every other region (400 × 400 matrix).

### Comparing functional connectivity and t-SNE analysis

To compare functional connectivity maps between the groups, we ran non-parametric permutation tests for each pairwise correlation value. This analysis was carried out twice: firstly, to compare edge differences between healthy individuals and Parkinson’s disease patients (control vs. PD), and secondly to compare edge differences between patients with and without visual hallucinations (PD+VH vs. PD-VH). From this, we created two binary matrices (control vs. PD; PD-VH vs. PD+VH), where values of 1 identified edges that were significantly different between the groups. To isolate edge differences uniquely associated with visual hallucinations, we subtracted the PD-VH vs. PD+VH binary matrix from the control vs. PD matrix. This resulted in a matrix with values of 1 representing edge differences only found between healthy controls and Parkinson’s disease, 0 representing edges different in both or none of the comparisons, and -1 for edges only different between patients with and without visual hallucinations. Edges with values of -1 were visualised on the cortical surface. These edge differences were unique to the comparison within the Parkinson’s disease cohort, consistent with differences specifically associated with visual hallucinations, rather than Parkinson’s disease in general. The same analysis was conducted on the t-SNE distance maps. Non-parametric tests were run for each correlation value (5,000 permutations),^55^ comparing control vs. PD, and PD-VH vs. PD+VH. The PD-VH matrix was subtracted from the PD+VH matrix isolating edge differences unique to the PD-VH vs. PD+VH comparison.

As a by-product of the above analysis, we noticed that group differences in the t-SNE distance maps appeared distinct from the group differences observed in the functional connectivity matrices. To determine whether the t-SNE distance maps did in fact describe distinct patterns, we ran eigendecomposition of the binary matrices into their eigenvectors and eigenvalues. Eigenvectors describe core patterns that underlie the high-dimensional binary matrix. The first eigenvector explains the most variance of the data and describes a principal pattern of group differences. The first eigenvector of the distance binary matrix for each comparison (control vs. PD, PD-VH vs. PD+VH) was then correlated with the corresponding first eigenvector of the correlation binary matrix to determine whether there was a common underlying pattern found in both the distance and correlation matrices.

### Comparing t-SNE results with the unimodal-heteromodal gradient

To determine whether the binary matrices described different patterns to those observed in the unimodal-to-heteromodal gradients, we averaged both binary matrices of the PD-VH vs. PD+VH comparison across regions. This resulted in a vector for each binary matrix that described the proportion of edges that were different between groups. We then correlated the edge vectors with the change in average gradient score across regions between Parkinson’s disease patients with and without visual hallucinations.

### Data availability

All code required to reproduce the statistical analyses and figures are publicly available (https://github.com/ShineLabUSYD/PD_Hallucinations).

## Results

### Demographic and clinical data

Demographic and clinical data was compared between patients and healthy controls, and within patients to compare hallucinators vs. non-hallucinators (Table 1). Sex ratio differed between the controls and overall patient group (*t* = 5.717, *p* = 0.017), but was equivalent in the hallucinating vs. non-hallucinating group (*t* = 0.589, *p* = 0.443). All the groups were matched for age and years of education, as well as for scores on the Montreal Cognitive Assessment, Logical Memory retention, and Trail-Making B-A score (*p* > 0.05). Performance in the Parkinson’s disease group was reduced relative to controls on some cognitive assessments, including the Mini-Mental State Examination (*t* = 2.962, *p* = 0.015), the Digit Span Task (*t* = 2.271, *p* = 0.033), and Part A of the Trail-Making test (*t* = 2.731, *p* = 0.016). The within-Parkinson’s groups were matched for daily dopamine dose (DDE), motor assessments (UPDRS-III, UPDRS-IV), Hoehn and Yahr scale, and the SCOPA-PC (*p* > 0.05). However, patients with visual hallucinations performed worse than non-hallucinating patients in Part B of the Trail-Making test (*t* = 2.305, *p* = 0.022), they reported a higher burden of daily motor problems (UDPRS-II; *t* = -3.836, *p* < 0.001), and they endorsed more severe mood symptoms on the Beck’s Depression Inventory (*t* = -2.302, *p* = 0.025) and the Hospital Anxiety and Depression Scale (*t* = -2.636, *p* = 0.011). There was a significant difference in total PsycH-Q scores, as patients with hallucinations had a higher burden of symptoms (*t* = 3.137, *p* = 0.004).

**Table 1.** Demographics. PD-VH = Parkinson’s disease without visual hallucinations. PD+VH = Parkinson’s disease with visual hallucinations. MMSE = Mini-Mental State Examination total score. MoCA = Montreal Cognitive Assessment total score. SCOPA-PC = Scales for Outcomes in Parkinson’s disease - Psychiatric Complications Total score. TMT A, TMT B are the z-scored trail-making test results. TMT B-A is the difference between TMT B and A and has been z-scored. BDI = Beck’s Depression Inventory total score. HADS = Hospital and Anxiety Depression Scale total score. UPDRS = Unified Parkinson’s Disease Rating Scale. Hoehn and Yahr total score. Sex comparisons were conducted using chi-square tests. All the continuous variables underwent pairwise comparisons with non-parametric permutation testing of the mean score. Significant p-values are in bold (*p* < 0.05). Section I of the UPDRS questionnaire as part of the questionnaire (Q2) was used to separate the patients into VH and nonVH groups.

|  | Control<br>(n = 19) |  |  | PD – VH<br>(n = 46) |  |  | PD + VH<br>(n = 31) |  |  | Control<br>vs<br>PD | PD – VH vs<br>PD + VH |
| --- | --- | --- | --- | --- | --- | --- | --- | --- | --- | --- | --- |
|  | n | Mean | SD | n | Mean | SD | n | Mean | SD | p-value | p-value |
| Gender<br>(Male:Female) | 8:11 |  |  | 36:10 |  |  | 21:10 |  |  | 0.017 | 0.443 |
| Age |  | 66.1 | 11.7 |  | 65.5 | 9.41 |  | 66.8 | 6.08 | 0.988 | 0.444 |
| Years of<br>Education |  | 14.1 | 2.76 |  | 14.6 | 2.95 |  | 13.7 | 3.38 | 0.755 | 0.234 |
| MMSE |  | 29.4 | 0.902 |  | 28.7 | 1.71 |  | 28.4 | 2.06 | **0.015** | 0.483 |
| MoCA |  | 27.6 | 2.27 |  | 27.1 | 2.95 |  | 26.5 | 2.95 | 0.238 | 0.326 |
| Digit Span |  | 12.7 | 2.77 |  | 10.9 | 2.84 |  | 11.5 | 2.91 | **0.033** | 0.404 |
| LM Retention |  | 12 | 2.52 |  | 11.6 | 2.88 |  | 11.5 | 3.25 | 0.539 | 0.930 |
| TMT A z-score |  | 0.684 | 0.884 |  | 0.251 | 0.823 |  | -0.289 | 1.35 | **0.016** | 0.053 |
| TMT B z-score |  | 0.663 | 0.918 |  | 0.133 | 0.851 |  | -0.591 | 1.61 | **0.005** | **0.022** |
| TMT B-A z-score |  | 1.05e-10 | 1.03 |  | -0.111 | 0.84 |  | -0.213 | 1.17 | 0.939 | 0.195 |
| BDI |  | 0.842 | 1.01 |  | 8.13 | 7.23 |  | 12.4 | 8.31 | **0.0001** | **0.025** |
| HADS |  | 3.74 | 3.54 |  | 6.13 | 4.77 |  | 10 | 7.17 | **0.002** | **0.011** |
| UPDRS |  |  |  |  |  |  |  |  |  |  |  |
| Section II |  |  |  |  | 8.26 | 6.41 |  | 15 | 8.25 |  | **0.0004** |
| Section III |  |  |  |  | 25.7 | 14.5 |  | 30.7 | 14.5 |  | 0.143 |
| Section IV |  |  |  |  | 0.804 | 1.87 |  | 1.65 | 2.99 |  | 0.172 |
| Hoehn and<br>Yahr |  |  |  |  | 1.93 | 0.574 |  | 2.15 | 0.503 |  | 0.096 |
| SCOPA-PC |  |  |  |  | 1.89 | 2.20 |  | 2.97 | 2.69 |  | 0.072 |
| DDE |  |  |  |  | 620 | 396 |  | 706 | 520 |  | 0.4511 |
| Psych-Q<br>(20 PD+VH, 27<br>PD-VH) |  |  |  |  | 5.85 | 5.87 |  | 13.95 | 10.11 |  | **0.004** |

### Functional connectivity nuisance variables

As expected, the Parkinson’s disease group as a whole had more head movements during scanning compared to controls, as indicated by higher framewise displacement (*t* = -3.486, *p* < 0.001), however there was no difference between patients with and without visual hallucinations (*t* = -1.414, *p* = 0.157). There was no significant correlation between participants’ head movement and average gradient score (*r* = 0.053, *p* = 0.607).

### Gradient connectivity analysis

The first gradient explained 14% of the variance and was anchored by the visual cortex at the lower end and the primary motor cortex at the upper end (Figure 2A). This gradient differed from the well-established unimodal-heteromodal gradient identified by Margulies and colleagues^22^ and it was not significantly correlated with the presumed brain network hierarchical organisation (*r* = 0.07, *p* = 0.16; Figure 2B).^22^ However, the second gradient, which explained 12% variance, did demonstrate a unimodal-heteromodal axis (Figure 2C) and was significantly correlated with network hierarchy organisation (*r* = 0.75; *p* < 0.05; Figure 2D). Therefore, we used the second gradient for our subsequent analyses concerned with unimodal-heteromodal organisation principles.

**Figure 1.**
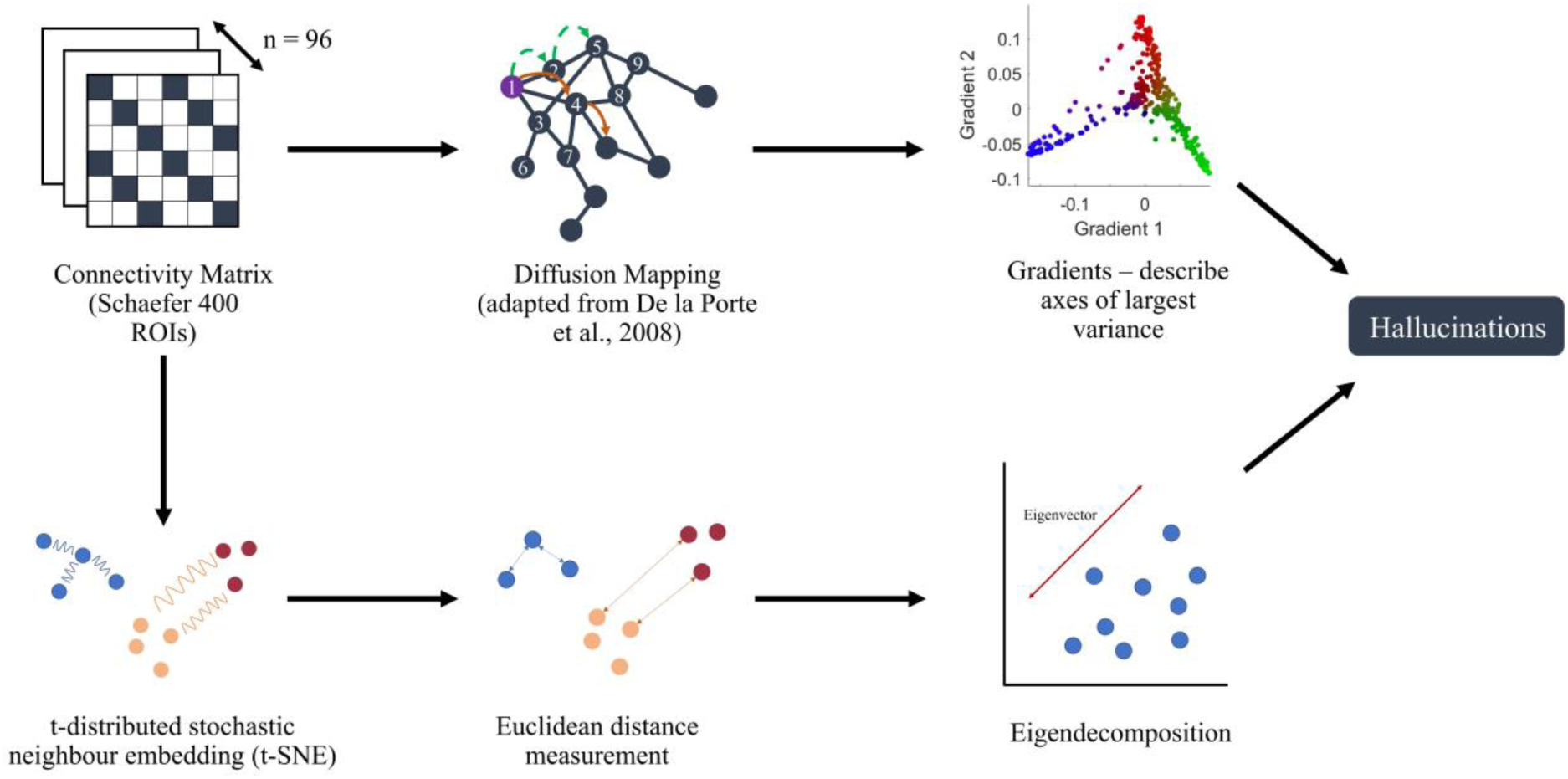
Summary of analyses conducted. A gradient map was constructed for each subject (*n* = 96) and group differences were analysed. A t-SNE map was also constructed for each subject (*n* = 96) and eigenvectors summarising the key differences between groups were defined.

**Figure 2.**
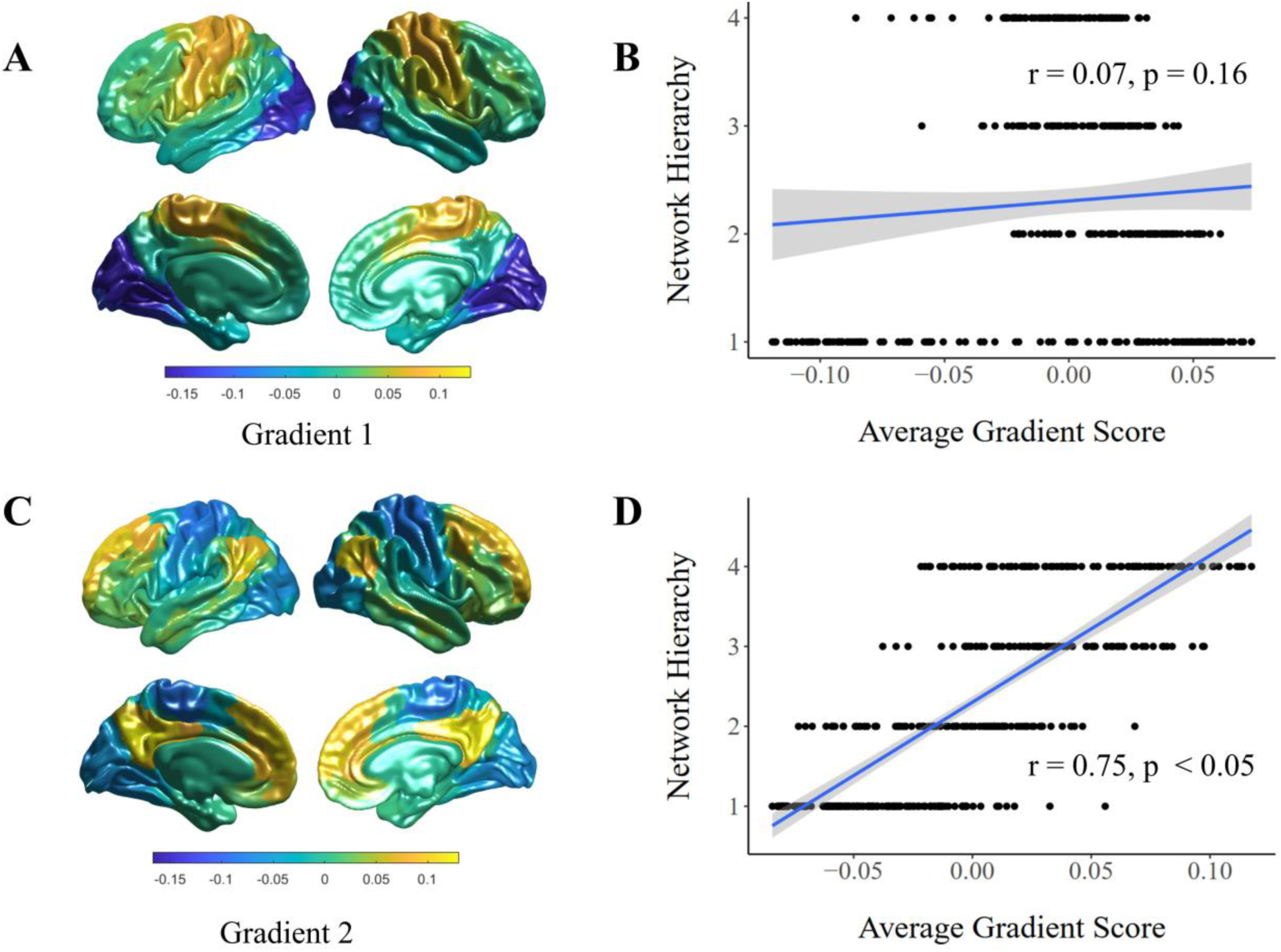
Comparison of gradients with network hierarchy organisation. **(A)** Population average of the first gradient that explains the most variance (14%). **(B)** First gradient score assigned to Yeo’s 7-network atlas and organised into proposed network hierarchy (1 = visual, somatomotor; 2 = dorsal attention, salience ventral attention; 3 = limbic, frontoparietal control; 4 = default mode network). **(C)** Population average of the second gradient that explains 12% variance. **(D)** Second gradient score assigned to Yeo’s 7-network atlas and organised into proposed network hierarchy.

Assessing the average gradient score distributions for each group (Figure 3A), all distributions were slightly right-skewed (*skewness* > 0), and they were also light-tailed – i.e., distribution of points were closer to the mean (*kurtosis* < 0). Distribution shape in the patient groups differed from controls (*D* = 0.057, *p_fdr_* < 0.05), but was not significantly different between the hallucinating and non-hallucinating patient groups (*D* = 0.015, *p_fdr_* = 0.06). However, visual comparison of the distributions suggests that the range of scores for the patients with visual hallucinations was smaller compared to the patients without visual hallucinations and the control group (Figure 3A).

**Figure 3.**
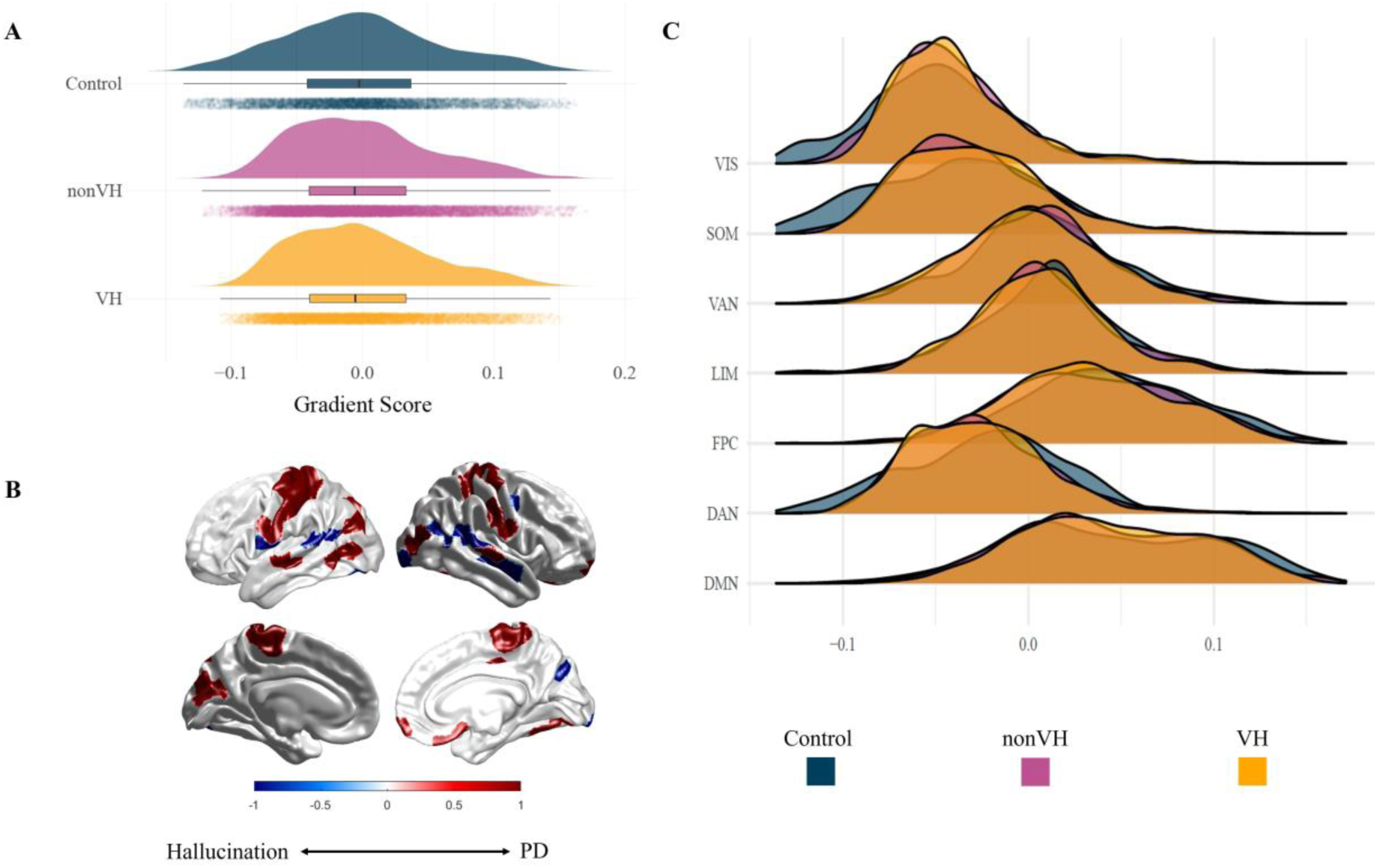
Group comparisons of gradient scores. **(A)** Distribution of average gradients score for each group. **(B)** Regions significantly different between groups. Values greater than 0 (red) refer to regions unique to Parkinson’s disease patients. Values less than 0 (blue) were regions unique to Parkinson’s disease patients with visual hallucinations. **(C)** Gradient score distributions across networks for each group.

### Group comparisons of gradient scores at the regional and network levels

Permutation testing was conducted at both the regional (*n* = 400) and network (*n* = 7) levels, comparing differences between Parkinson’s disease patients against controls, and differences between the Parkinson’s disease groups. These results are shown in Figure 3B. Prominent differences between the patients and controls were observed in regions of the primary motor cortex (*p* < 0.05); differences were also found in regions from the extra-striate visual cortex and laterally in the temporal lobe (*p* < 0.05). Regions significantly different for patients with visual hallucinations consisted of regions near the temporoparietal junction (*p* < 0.05). For all these regions, the gradient score was higher in the disease groups.

By assigning each of the 400 regions to the 7-network atlas,^51^ we observed group differences at the network level (Figure 3C). Comparing controls and Parkinson’s disease patients, the average gradient score significantly increased in patients for the visual and somatomotor networks (*p* < 0.05, *mean difference* = 0.007 and 0.008, respectively). In contrast, there were significant decreases in patients’ gradient scores for the ventral attentional and frontoparietal control networks (*p* < 0.05, *mean difference* = -0.005 and -0.009, respectively). In Parkinson’s disease patients with versus without visual hallucinations, significant gradient score differences were observed in the visual, ventral attention and frontoparietal control networks (*p* < 0.05). Patients with visual hallucinations had higher gradient scores in the visual network (*mean difference* = 0.003), and lower gradient scores in the ventral attention (*mean difference* = - 0.005) and frontoparietal control networks (*mean difference* = -0.003). Overall, these results demonstrated reduced functional separation between sensory and higher-order networks along the unimodal-heteromodal axis in patients with visual hallucinations.

### Relationship between average network gradient score, TMT-B, BDI and PsycH-Q

Average gradient scores of the visual, ventral attention, and frontoparietal control networks were compared against clinical scores that differed significantly between the groups. The ventral attention network average gradient score was significantly correlated to performance in the TMT-B and BDI (*r* = 0.210, *p* = 0.040; *r* = -0.228, *p* = 0.026, respectively), consistent with impaired performance on both measures being associated with gradient scores shifted towards the sensory regions. However, these results did not survive false discovery rate correction (*p_fdr_* > 0.05). No other comparisons between these clinical measures and network gradient scores were significant (*p* > 0.05).

### Group differences in functional connectivity

Functional connectivity comparisons between the Parkinson’s disease and control groups showed increases within the somatomotor network and in edges connecting the somatomotor network with the visual, dorsal attention and default mode networks (*p* < 0.05). Specifically, these were edges between the primary motor cortex to the extra-striate cortex, parietal lobe, and regions of the temporoparietal junction. There was also a significant increase in edges connecting regions of the somatomotor network with dorsal and lateral regions of the prefrontal cortex (*p* < 0.05). Comparing Parkinson’s disease patients with and without visual hallucinations revealed a distinct pattern of increased connectivity in edges between the visual and default mode networks (*p* < 0.05). These included regions in the temporal lobe, temporoparietal junction, extra-striate cortex. There was also a secondary pattern involving edges between the primary motor cortex, superior parietal lobe and the frontal lobe (*p* < 0.05).

To focus on differences that might relate specifically to visual hallucinations, we looked at the edges that differed between patients with and without hallucinations but did not differ when comparing the patient group as a whole with controls. These edges were also unique to the correlation matrix and were not significantly different in the Euclidean matrix from the t-SNE results. Overall, edges between the visual and default mode networks, specifically regions of the extra-striate cortex, temporal, parietal, and frontal lobes were unique to Parkinson’s disease patients with visual hallucinations (Figure 5A).

### Group differences in t-SNE distance analysis

Figure 4 shows the t-SNE embedding from each group. Between Parkinson’s disease and healthy controls, a significant increase in Euclidean distance was found in the t-SNE analysis for the limbic, ventral attention, and executive (frontoparietal control, default mode) networks (*p* < 0.05). Specifically, these regions were from the inferior parietal lobule, medial regions of the motor cortex, ventral and lateral prefrontal cortex, and the inferior and superior regions of the temporal lobe. These reconfigurations included somatomotor regions that have been assigned to the visual network of the Yeo 7-network atlas. There was also an increase in distance between the extra-striate cortex and the rest of the visual network (*p* < 0.05).

**Figure 4.**
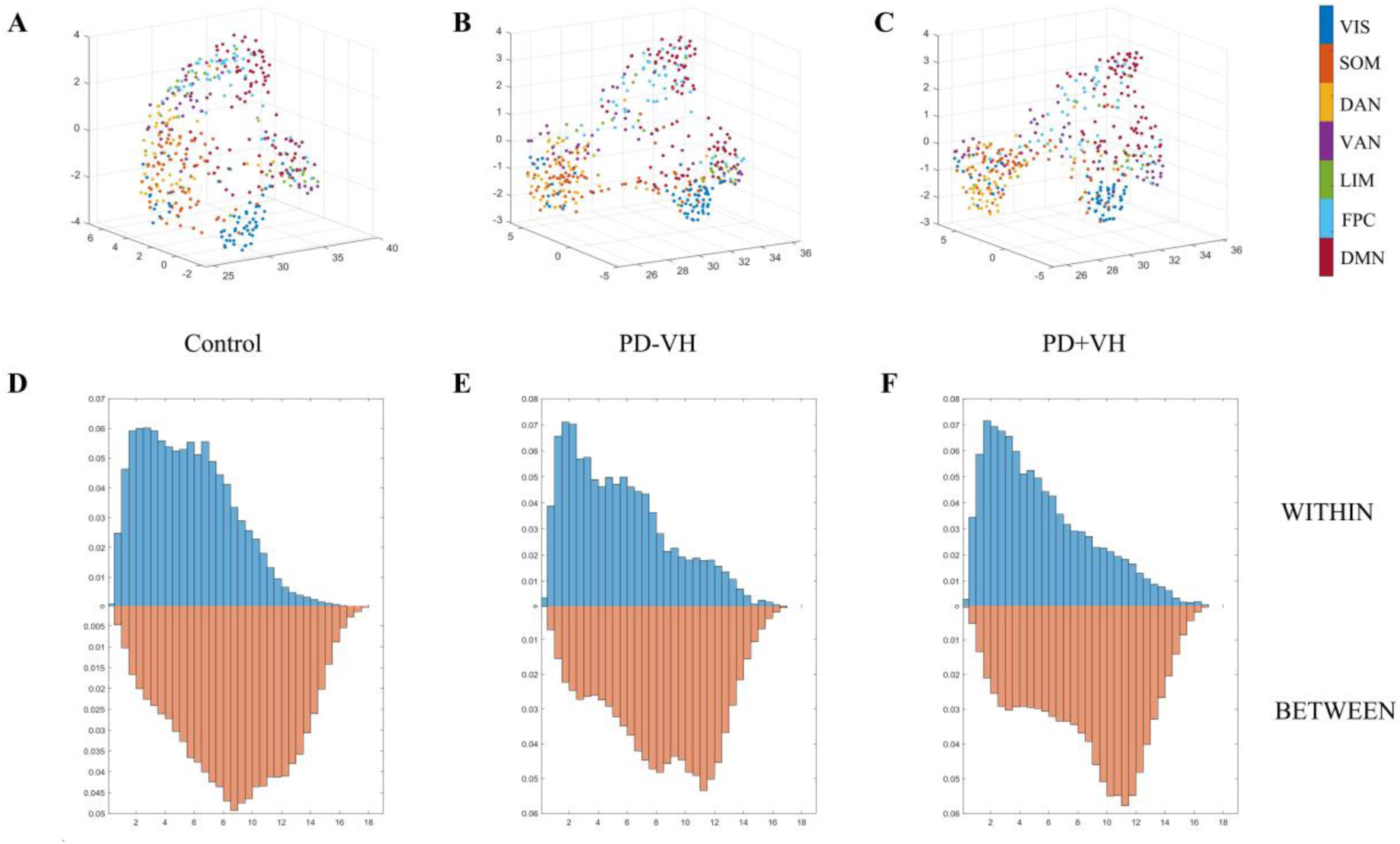
t-SNE plots of 400 Schaefer regions. **(A)** t-SNE plot of control group average functional connectivity. **(B)** t-SNE plot of PD-VH average functional connectivity. **(C)** t-SNE plot of PD+VH average functional connectivity. Plots A-C are coloured by Yeo 7-network atlas. **(D-F)** Density histograms of the Euclidean distance within vs. between networks for each t-SNE plot.

Narrowing the focus to differences between patients with and without visual hallucinations, we found increased Euclidean distances from the visual network to the superior and inferior regions of the parietal cortex, lateral and ventral regions of the prefrontal cortex, and the posterior cingulate (*p* < 0.05). There was also a significant increase in distance between regions of the lateral motor cortex and temporal occipital cortex to regions of the frontoparietal control and default mode networks (*p* < 0.05). Significant increases in distance in the superior temporal lobe and the temporal pole of the right hemisphere were also evident (*p* < 0.05).

From the t-SNE results, we isolated differences that were unique to patients with visual hallucinations. Patients with visual hallucinations had an increased Euclidean distance between regions from the dorsal and ventral prefrontal cortex, lateral motor cortex, inferior parietal cortex, temporal occipital cortex, and posterior cingulate. The unilateral increased distances found in the right temporal pole and superior temporal cortex were also attributed to patients with visual hallucinations only (Figure 5B). Overall, the t-SNE results showed that brain regions were situated further apart (less compressed) in patients with visual hallucinations.

**Figure 5.**
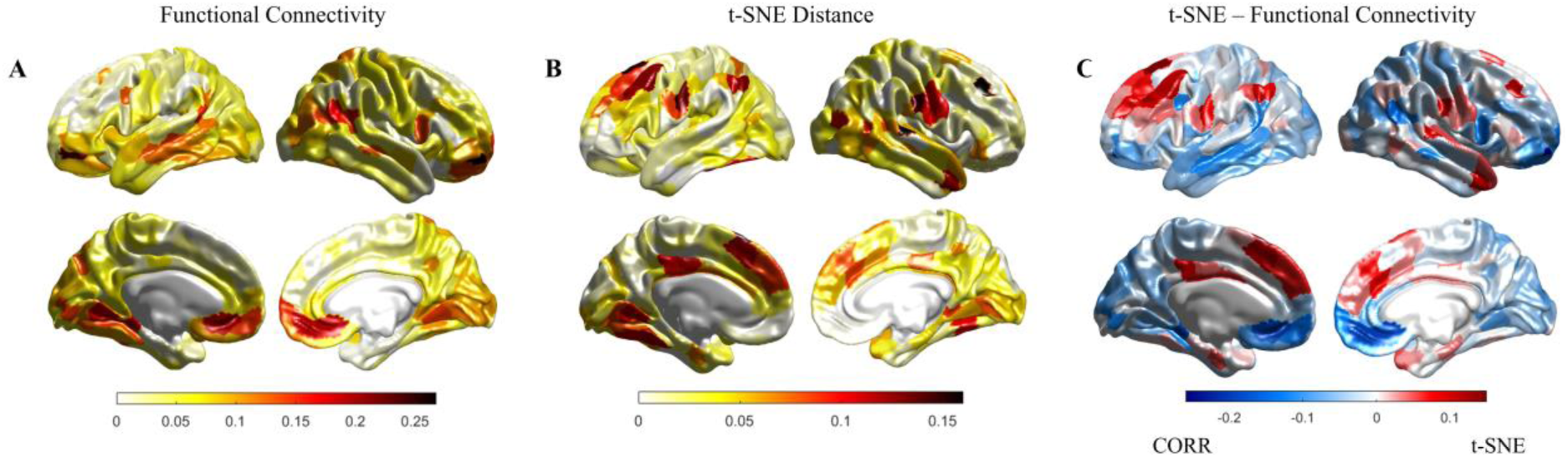
Proportion of edges that defined patients with hallucinations from the other two groups. **(A)** Proportion of edges that were different in functional connectivity. **(B)** Proportion of edges that were different in Euclidean distance. **(C)** Proportion of edges that are different in Euclidean distance compared to functional connectivity. Regions coloured in red have more differences in Euclidean distance, and regions coloured in blue have more differences in functional connectivity.

### Relationship between functional connectivity and t-SNE distances

Group differences observed by analysing the correlation matrix were not always replicated in the t-SNE Euclidean distance matrix. For instance, comparing the correlation and t-SNE differences for the control vs. Parkinson’s disease groups showed that increased functional connectivity in the primary motor cortex was mirrored by increased Euclidean distance in the t-SNE analysis. However, this contradicts our intuition of the relationship between functional connectivity and Euclidean distance: an *increase* in functional connectivity should equate to a *decrease* in Euclidean distance. Using this intuition as a guide, we could investigate the latent network signatures. In doing so, we noticed that the prominent between-group differences in standard functional connectivity between the visual and somatomotor networks were not upheld by the t-SNE analysis. Specifically, the increased Euclidean distance between the somatomotor network and higher order networks we observed on the t-SNE plots were not as prominent in the correlation matrix, suggesting that passing the functional data through the unique filter of the t-SNE was sufficient to expose specific differences in network-level organisation that were not detectable through standard functional connectivity analyses.

Given the difference in interpretation associated with the functional connectivity and t-SNE matrices, we compared the two directly – i.e., comparing the core patterns of the binary matrices by finding the eigenvectors of the correlation and t-SNE Euclidean distance matrices through eigendecomposition. This can be interpreted as capturing latent components of the original low-dimensional embedding: the first eigenvector that explains the most variance of the data describes a pattern that differentiates the Parkinson’s disease group from controls. In the first eigenvector for functional connectivity, the main differences between the controls vs. Parkinson’s disease were in regions of the occipital lobe, temporal and frontal pole, motor cortex and anterior cingulate (Figure 6A). For the first eigenvector for Euclidean distance, main differences were observed in the prefrontal cortex, inferior temporal cortex and temporal parietal junction laterally, both posterior and anterior cingulate cortex medially, primary motor cortex, extra-striate cortex, and regions of the superior temporal cortex (Figure 6C). We then established whether these patterns were equivalent by calculating the Pearson’s correlation between the two eigenvectors. The two eigenvectors were only weakly correlated (*r* = 0.093, *p* = 0.065), confirming that the correlation and Euclidean distance matrices highlight distinct differences between the control and Parkinson’s disease groups.

**Figure 6.**
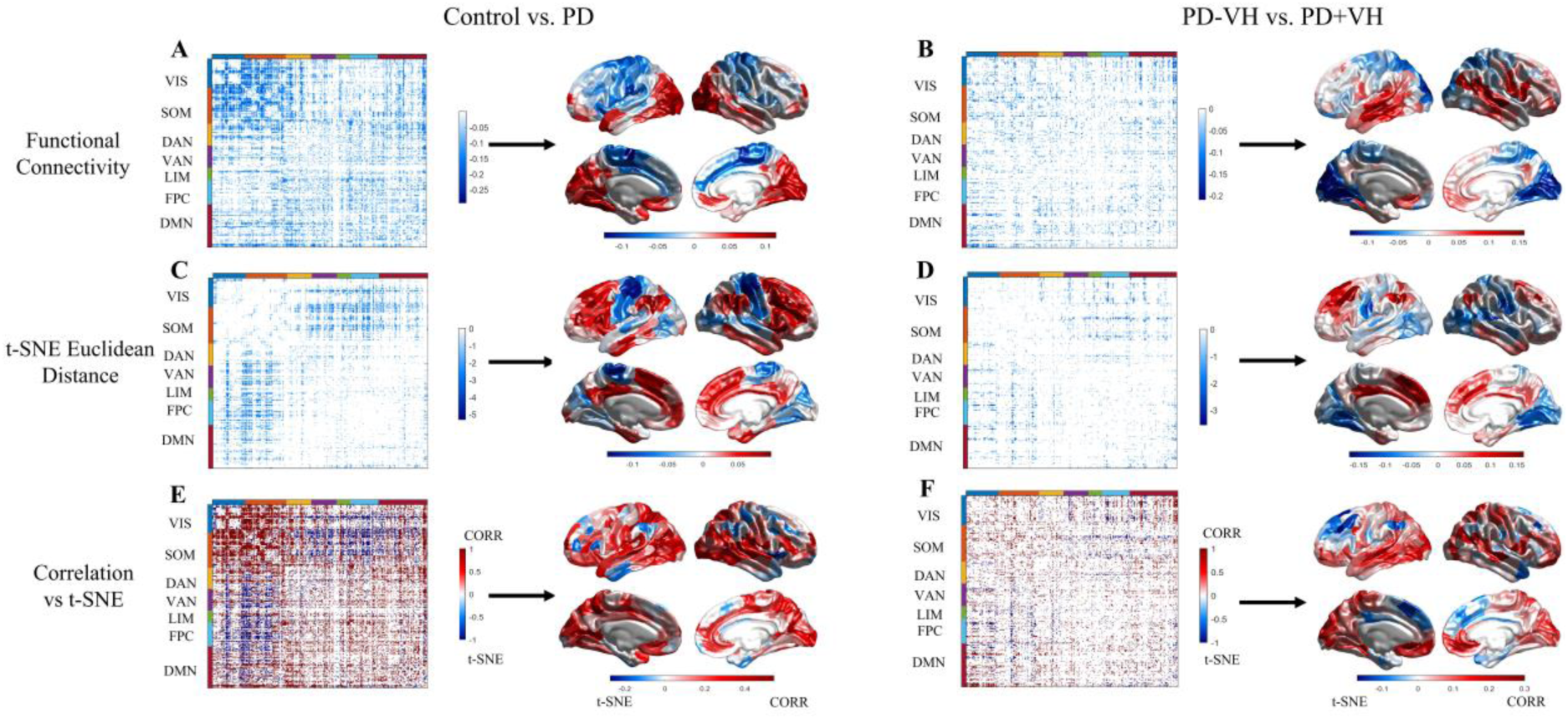
Functional connectivity (FC) and t-SNE analysis for Control vs Parkinson’s disease. Left column **(A, C, E)** refers to Control vs. PD comparisons. Right column **(B, D, F)** refers to PD-VH vs. PD+VH comparisons. The first row **(A, B)** were group differences in FC. Matrices were coloured by group differences (Control - PD; PD-VH - PD+VH respectively), with values less than 0 indicating higher FC between regions in PD and PD+VH respectively. The first eigenvector of each matrix was visualised on the cortical surface. The second row **(C, D)** refers to group differences in Euclidean distance between t-SNE maps. Matrices were coloured by group differences (Control - PD; PD-VH - PD+VH respectively), with values less than 0 indicating greater Euclidean distance between regions in PD and PD+VH respectively. The first eigenvector of each matrix was visualised on the cortical surface. The third row **(E, F)** refers to differences between the FC and t-SNE results. Matrices were coloured by group differences, with 1 indicating a difference observed from FC analysis, -1 indicating a difference observed from t-SNE analysis, and 0 indicating a difference observed in either both or none of the analyses. The average difference between analyses across regions was visualised on the cortical surface such that *values* > 0 indicate differences more common in FC analysis, and *values* < 0 indicate differences more common in t-SNE analysis.

Comparing patients with and without visual hallucinations, there were no edge differences that overlapped between the correlation and Euclidean distance matrices. Similar to the previous comparison, we can confirm that these matrices describe different patterns by calculating the Pearson’s correlation between the eigenvector for each matrix. For the correlation eigenvector, main differences were found in the temporal lobe, extending to the temporoparietal junction, the primary visual cortex, the medial extrastriate cortex and parts of the parietal cortex (Figure 6B). For the Euclidean distance eigenvector, the main differences were in the prefrontal cortex, inferior parietal cortex, lateral motor cortex and regions of the extrastriate cortex (Figure 6D). The two eigenvectors were weakly correlated (*r* = 0.063, *p* = 0.209), confirming that the correlation and Euclidean distance matrices differentiated between patients with and without visual hallucinations in distinct ways. In summary, the functional connectivity correlation matrices revealed increased connectivity between primary visual and temporal regions, whereas the t-SNE matrices showed increased Euclidean distances between the prefrontal, motor and extrastriate cortex.

### Relationship between t-SNE distances and the unimodal-heteromodal gradient

Group differences found in the correlation and t-SNE analyses were compared against differences observed in the unimodal-heteromodal gradients. We did this in two steps: 1) by identifying the proportion of edges that were significantly different between groups for each region in the correlation and Euclidean distance matrices; and 2) we then compared whether the number of edges that were different between the groups correlated with the change in gradient scores between patients with and without visual hallucinations. Pearson’s correlation confirmed a weak negative correlation between the proportion of edges in the Euclidean matrix and the change in gradient score (*r* = -0.100, *p* = 0.05). For the proportion of edges in the correlation matrix, there was a strong negative correlation with the change in gradient scores (*r* = -0.324, *p* < 0.001). Therefore, the correlation matrix differences aligned more with differences in gradient scores compared to the t-SNE results, further confirming that the t-SNE analysis revealed unique signatures from patients with visual hallucinations that are not found when directly interrogating the raw functional correlation matrix.

## Discussion

Here we combined insights across functional connectivity, cortical gradients and t-SNE distance mapping, to demonstrate the altered network hierarchy in Parkinson’s disease visual hallucinations. In patients with hallucinations, gradients analysis revealed increased functional integration (i.e., compression) between sensory and higher order networks. This was mirrored by results from the correlation matrices, which showed increased connectivity between the visual and default-mode networks in the hallucinating group. However, when projecting into the t-SNE space, new reconfigurations that defined hallucinating patients were revealed. The hallucinating group was characterised by increased Euclidean distances in edges connecting regions of the visual network to the frontoparietal control and default mode networks, as well as edges within the default mode network. Furthermore, group differences observed in the t-SNE space were only weakly correlated with differences in the unimodal-heteromodal functional gradient, compared to the strong correspondence between the correlation matrix and gradient results. Together, our results confirm altered network hierarchy in Parkinson’s disease hallucinations across multiple dimensionality reduction techniques. Furthermore, our novel application of t-SNE distance analysis may provide new insights into the neural signatures of visual hallucinations – exposing non-linear, network-level reconfigurations not typically identifiable in traditional functional connectivity and gradient analyses.

Compression of the unimodal-heteromodal gradient may disrupt perceptual processing and contribute to hallucination vulnerability. Separation between functional regions in the gradient context has been linked to spatial separation along the cortex, with long-range connections between unimodal and heteromodal regions serving as one of the foundations for information processing.^56, 57^ In Parkinson’s disease with visual hallucinations, there was increased functional integration between sensory and higher order networks, as gradient scores for regions from the visual, attentional, and frontoparietal control networks shifted closer together on the gradient. Decreased separation along the gradient implies increased similarity in connectivity profiles and increased integration between regions. Indeed, we observed higher functional connectivity between the visual network and regions of the ventral attention, frontoparietal control, and default mode networks in patients with visual hallucinations. This is consistent with previous work showing increased coupling between sensory and higher-order networks in Parkinson’s disease and other neuropsychiatric disorders that involve perceptual disturbances.^13, 14, 58, 59^ Increased connectivity between higher-order and sensory regions, and compression of the unimodal-heteromodal gradient, are routes that may permit excessive influence over earlier perceptual processes – consistent with the proposal that abnormal modulation from top-down regions over the visual system increases susceptibility to visual hallucinations.^1, 8, 60–63^

Compression of the unimodal-heteromodal gradient is not unique to Parkinson’s disease and may be a transdiagnostic feature in disorders with perceptual disturbances. Previous studies in other neuropsychiatric disorders, including autism spectrum disorder and schizophrenia, also showed compression of the unimodal-heteromodal gradient.^24, 25^ Both studies observed an association between changes in the gradient and measures of disease severity.^24, 25^ Similarly, in Parkinson’s disease, the extent of visual impairment has been related to the amount of compression in the unimodal-heteromodal gradient.^26^ We observed an association between gradient scores and measures of attentional set-shifting and mood – consistent with poorer attention and increased mood symptoms associated with a lower (more compressed) gradient score. While these are not direct clinical measures of hallucination severity, attention and mood problems are prominent features of the hallucinating phenotype^64, 65^ and may predict the development of visual hallucinations.^66^ Taken together, compression of the unimodal-heteromodal gradient has been observed across neuropsychiatric disorders, varying with clinical measures of disease severity, and may be a predisposing trait for hallucinations in Parkinson’s disease.

Between the groups, there was increased functional differentiation between sensory and higher-order networks in the t-SNE space. The distance between regions in the t-SNE plot are based on the similarity of their functional connectivity profiles,^29^ with regions functionally similar to each other placed closer together in t-SNE space. From this intuition, regions that share increased functional connectivity should be mirrored by decreased Euclidean distance (and *vice versa*). However, we observed certain regions with relatively strong functional correlations in hallucinating patients were increasingly separated in t-SNE space. Regional pairs that demonstrate this pattern may have strong correlations but retain the ability for relatively segregated processing, based on the highly non-linear patterns stored within the rest of the network. Importantly, most regional pairs we observed with this pattern were in the prefrontal cortex, which serves an important role in “top-down” perceptual processing,^67, 68^ possibly via its initial rapid processing of ambiguous information via magnocellular pathways.^69, 70^ Abnormalities in the prefrontal cortex reduce cognitive flexibility and impair performance in tasks that involve distractions or ambiguous information.^67^ Increased distance between the prefrontal cortex and sensory regions suggests reduced interactions and functional coupling between these regions, possibly decreasing the reliance on the prefrontal cortex for information processing, and impairing the ability to process ambiguous information. These results demonstrate that the t-SNE analysis captures a unique facet of hallucination susceptibility, which is complementary to the functional gradients results and not typically identified in correlation matrix analyses.

The current study focused on neuroimaging analyses of patients at rest, however previous studies measuring hallucination-like events in Parkinson’s disease have converged on similar brain regions, suggesting these regions are functionally relevant to the hallucination state.^8^ Perception is influenced by past experiences, reconciling incoming sensory information with known statistics about the external world – allowing us to predict and interpret incoming information, even in ambiguous situations.^1, 2^ From resting state analyses, we observed reconfigurations in network organisation that could disrupt how our perceptual system processes internal and external information. Compression of the unimodal-heteromodal gradient suggests that in patients susceptible to visual hallucinations, there is potential for increased influence from top-down processes to override sensory information. The t-SNE analysis highlights increased differentiation between the prefrontal cortex and sensory regions, which may result in a decreased reliance on the prefrontal cortex for processing ambiguous information. Taken together, when patients vulnerable to hallucinations encounter environments with minimal sensory information, they may be unable to process the ambiguous information appropriately and increase their dependence on internal associations, resulting in misleading predictions of their surrounding environment and the formation of hallucinations.^2, 63^

Treatment of visual hallucinations and psychosis in Parkinson’s disease is challenging, and dimensionality reduction techniques may provide a novel objective for medicinal drugs.^71^ The standard dopaminergic medication has minimal benefits on hallucinations, and can in some cases exacerbate them.^72, 73^ However, treatment with classic antipsychotic medications can have adverse secondary effects, including worsened motor symptoms.^74, 75^ A newer medication for Parkinson’s disease psychosis is the selective serotonin inverse agonist pimavanserin, which acts primarily at the 5-HT_2A_ receptor.^71, 76^ This drug, which effectively antagonises the 5-HT_2A_ receptor, improves psychosis symptoms without the unintended motor side effects.^74, 76^ Single-dose studies in healthy people using pro-serotonergic drugs (acting primarily at 5-HT_2A_) reveal a compression of the principal gradient induced by drugs that agonise 5-HT_2A_.^77^ It could be speculated that pimavaserin may promote a recovery of separation within the principal gradient, consistent with a role for 5-HT_2A_ activity in modulating the degree of feedback and information transfer in the brain.^78^ This opens the possibility of gradient analysis being a useful means for predicting and measuring the beneficial effects of drug treatments for visual hallucinations – consistent with the broader goal of establishing neuroimaging signatures that can advance personalised drug treatment in Parkinson’s disease.^79^

This study revealed reconfigurations in network interactions for Parkinson’s disease patients susceptible to visual hallucinations. Compression of the unimodal-heteromodal gradient was associated with cognitive performance and may be a useful measurement for understanding the extent of abnormal interactions between top-down and bottom-up processing. Furthermore, this study demonstrated that projecting functional connectivity into the t-SNE space provides an alternate perspective to hallucinations in Parkinson’s disease that is overlooked in traditional functional connectivity analyses. With continued advancements in imaging methods and increased diversity in neuropsychiatric data, dimensionality reduction techniques are a lens through which the neural signatures of hallucinations, across modalities and disorders, might be reconciled.

## References

1. Collerton D, Perry E, McKeith I. Why people see things that are not there: A novel Perception and Attention Deficit model for recurrent complex visual hallucinations. Behav Brain Sci. 2005;28(6):737–757. doi:10.1017/S0140525X05000130

2. Hardstone R, Zhu M, Flinker A, et al. Long-term priors influence visual perception through recruitment of long-range feedback. Nat Commun. 2021;12(1):6288. doi:10.1038/s41467-021-26544-w

3. Gilbert CD, Li W. Top-down influences on visual processing. Nat Rev Neurosci. 2013;14(5):350–363. doi:10.1038/nrn3476

4. Powers AR, Kelley M, Corlett PR. Hallucinations as Top-Down Effects on Perception. Biological Psychiatry: Cognitive Neuroscience and Neuroimaging. 2016;1(5):393–400. doi:10.1016/j.bpsc.2016.04.003

5. O’Callaghan C, Kveraga K, Shine JM, Adams RB, Bar M. Predictions penetrate perception: Converging insights from brain, behaviour and disorder. Consciousness and Cognition. 2017;47:63–74. doi:10.1016/j.concog.2016.05.003

6. Thomas GE, Zeidman P, Sultana T, Zarkali A, Razi A, Weil RS. Changes in Both Top-down and Bottom-up Effective Connectivity Drive Visual Hallucinations in Parkinson’s Disease. Neuroscience; 2022. doi:10.1101/2022.03.25.485621

7. Shine JM, O’Callaghan C, Halliday GM, Lewis SJG. Tricks of the mind: Visual hallucinations as disorders of attention. Progress in Neurobiology. 2014;116:58–65. doi:10.1016/j.pneurobio.2014.01.004

8. Shine JM, Keogh R, O’Callaghan C, Muller AJ, Lewis SJG, Pearson J. Imagine that: elevated sensory strength of mental imagery in individuals with Parkinson’s disease and visual hallucinations. Proc R Soc B. 2015;282(1798):20142047. doi:10.1098/rspb.2014.2047

9. O’Callaghan C, Firbank M, Tomassini A, Schumacher J, O’Brien JT, Taylor JP. Impaired sensory evidence accumulation and network function in Lewy body dementia. Brain Communications. 2021;3(3):fcab089. doi:10.1093/braincomms/fcab089

10. Rollins CPE, Garrison JR, Simons JS, et al. Meta-analytic Evidence for the Plurality of Mechanisms in Transdiagnostic Structural MRI Studies of Hallucination Status. EClinicalMedicine. 2019;8:57–71. doi:10.1016/j.eclinm.2019.01.012

11. Okuneye VT, Meda S, Pearlson GD, et al. Resting state auditory-language cortex connectivity is associated with hallucinations in clinical and biological subtypes of psychotic disorders. NeuroImage: Clinical. 2020;27:102358. doi:10.1016/j.nicl.2020.102358

12. Spinosa V, Brattico E, Campo F, Logroscino G. A systematic review on resting state functional connectivity in patients with neurodegenerative disease and hallucinations. NeuroImage: Clinical. 2022;35:103112. doi:10.1016/j.nicl.2022.103112

13. Shine JM, Muller AJ, O’Callaghan C, Hornberger M, Halliday GM, Lewis SJ. Abnormal connectivity between the default mode and the visual system underlies the manifestation of visual hallucinations in Parkinson’s disease: a task-based fMRI study. npj Parkinson’s Disease. 2015;1(1):15003. doi:10.1038/npjparkd.2015.3

14. Walpola IC, Muller AJ, Hall JM, et al. Mind-wandering in Parkinson’s disease hallucinations reflects primary visual and default network coupling. Cortex. 2020;125:233–245. doi:10.1016/j.cortex.2019.12.023

15. Cunningham JP, Yu BM. Dimensionality reduction for large-scale neural recordings. Nat Neurosci. 2014;17(11):1500–1509. doi:10.1038/nn.3776

16. Shine JM, Hearne LJ, Breakspear M, et al. The Low-Dimensional Neural Architecture of Cognitive Complexity Is Related to Activity in Medial Thalamic Nuclei. Neuron. 2019;104(5):849–855.e3. doi:10.1016/j.neuron.2019.09.002

17. Vos de Wael R, Benkarim O, Paquola C, et al. BrainSpace: a toolbox for the analysis of macroscale gradients in neuroimaging and connectomics datasets. Commun Biol. 2020;3(1):103. doi:10.1038/s42003-020-0794-7

18. Coifman RR, Lafon S, Lee AB, et al. Geometric diffusions as a tool for harmonic analysis and structure definition of data: Diffusion maps. Proc Natl Acad Sci USA. 2005;102(21):7426–7431. doi:10.1073/pnas.0500334102

19. Langs G, Golland P, Ghosh SS. Predicting Activation Across Individuals with Resting-State Functional Connectivity Based Multi-Atlas Label Fusion. Med Image Comput Comput Assist Interv. 2015;9350:313–320. doi:10.1007/978-3-319-24571-3_38

20. Haak KV, Marquand AF, Beckmann CF. Connectopic mapping with resting-state fMRI. NeuroImage. 2018;170:83–94. doi:10.1016/j.neuroimage.2017.06.075

21. Huntenburg JM, Bazin PL, Margulies DS. Large-Scale Gradients in Human Cortical Organization. Trends in Cognitive Sciences. 2018;22(1):21–31. doi:10.1016/j.tics.2017.11.002

22. Margulies DS, Ghosh SS, Goulas A, et al. Situating the default-mode network along a principal gradient of macroscale cortical organization. Proc Natl Acad Sci USA. 2016;113(44):12574–12579. doi:10.1073/pnas.1608282113

23. Bethlehem RAI, Paquola C, Seidlitz J, et al. Dispersion of functional gradients across the adult lifespan. NeuroImage. 2020;222:117299. doi:10.1016/j.neuroimage.2020.117299

24. Hong SJ, Vos de Wael R, Bethlehem RAI, et al. Atypical functional connectome hierarchy in autism. Nat Commun. 2019;10(1):1022. doi:10.1038/s41467-019-08944-1

25. Dong D, Yao D, Wang Y, et al. Compressed sensorimotor-to-transmodal hierarchical organization in schizophrenia. Psychol Med. Published online June 8, 2021:1–14. doi:10.1017/S0033291721002129

26. Zarkali A, McColgan P, Leyland LA, Lees AJ, Rees G, Weil RS. Organisational and neuromodulatory underpinnings of structural-functional connectivity decoupling in patients with Parkinson’s disease. Commun Biol. 2021;4(1):86. doi:10.1038/s42003-020-01622-9

27. Kobak D, Berens P. The art of using t-SNE for single-cell transcriptomics. Nat Commun. 2019;10(1):5416. doi:10.1038/s41467-019-13056-x

28. Böhm JN, Berens P, Kobak D. Attraction-Repulsion Spectrum in Neighbor Embeddings. Published online December 3, 2021. Accessed May 15, 2022. http://arxiv.org/abs/2007.08902

29. van der Maaten L. Barnes-Hut-SNE. Published online March 8, 2013. Accessed May 17, 2022. http://arxiv.org/abs/1301.3342

30. 30. Arora S, Hu W, Kothari PK. An Analysis of the t-SNE Algorithm for Data Visualization. In: Proceedings of the 31st Conference On Learning Theory. PMLR; 2018:1455–1462. Accessed May 16, 2022. https://proceedings.mlr.press/v75/arora18a.html

31. Belkina AC, Ciccolella CO, Anno R, Halpert R, Spidlen J, Snyder-Cappione JE. Automated optimized parameters for T-distributed stochastic neighbor embedding improve visualization and analysis of large datasets. Nat Commun. 2019;10(1):5415. doi:10.1038/s41467-019-13055-y

32. Martinez-Martin P, Falup-Pecurariu C, Rodriguez-Blazquez C, et al. Dementia associated with Parkinson’s disease: Applying the Movement Disorder Society Task Force criteria. Parkinsonism & Related Disorders. 2011;17(8):621–624. doi:10.1016/j.parkreldis.2011.05.017

33. Goetz CG, Tilley BC, Shaftman SR, et al. Movement Disorder Society-sponsored revision of the Unified Parkinson’s Disease Rating Scale (MDS-UPDRS): Scale presentation and clinimetric testing results. Movement Disorders. 2008;23(15):2129–2170. doi:https://doi.org/10.1002/mds.22340

34. Tomlinson CL, Stowe R, Patel S, Rick C, Gray R, Clarke CE. Systematic review of levodopa dose equivalency reporting in Parkinson’s disease. Movement Disorders. 2010;25(15):2649–2653. doi:10.1002/mds.23429

35. Visser M, Verbaan D, Rooden SM van, Stiggelbout AM, Marinus J, Hilten JJ van. Assessment of psychiatric complications in Parkinson’s disease: The SCOPA-PC. Movement Disorders. 2007;22(15):2221–2228. doi:https://doi.org/10.1002/mds.21696

36. Shine JM, Mills JMZ, Qiu J, et al. Validation of the Psychosis and Hallucinations Questionnaire in Non-demented Patients with Parkinson’s Disease. Mov Disord Clin Pract. 2015;2(2):175–181. doi:10.1002/mdc3.12139

37. Folstein MF. The Mini-Mental State Examination. Arch Gen Psychiatry. 1983;40(7):812. doi:10.1001/archpsyc.1983.01790060110016

38. Nasreddine ZS, Phillips NA, BÃ©dirian V, et al. The Montreal Cognitive Assessment, MoCA: A Brief Screening Tool For Mild Cognitive Impairment: MOCA: A BRIEF SCREENING TOOL FOR MCI. Journal of the American Geriatrics Society. 2005;53(4):695–699. doi:10.1111/j.1532-5415.2005.53221.x

39. Bowie CR, Harvey PD. Administration and interpretation of the Trail Making Test. Nat Protoc. 2006;1(5):2277–2281. doi:10.1038/nprot.2006.390

40. Wechsler D. Wechsler Adult Intelligence Scale--Third Edition. Published online 1977. doi:https://doi.org/10.1037/t49755-000

41. R Core Team. R: A language and environment for statistical computing. R Foundation for Statistical Computing, Vienna, Austria. Published online 2022. https://www.R-project.org/

42. van Buuren S. Multivariate Imputation by Chained Equations. Published online November 23, 2021. Accessed October 19, 2022. https://cran.r-project.org/web/packages/mice/mice.pdf

43. Gorgolewski KJ, Auer T, Calhoun VD, et al. The brain imaging data structure, a format for organizing and describing outputs of neuroimaging experiments. Sci Data. 2016;3(1):160044. doi:10.1038/sdata.2016.44

44. 44. Li X. DICOM to NIfTI conversion, DICOM and NIfTI tools, NIfTI visualization. Published online 2022. https://github.com/xiangruili/dicm2nii

45. Li X, Morgan PS, Ashburner J, Smith J, Rorden C. The first step for neuroimaging data analysis: DICOM to NIfTI conversion. Journal of Neuroscience Methods. 2016;264:47–56. doi:10.1016/j.jneumeth.2016.03.001

46. Esteban O, Markiewicz CJ, Blair RW, et al. fMRIPrep: a robust preprocessing pipeline for functional MRI. Nat Methods. 2019;16(1):111–116. doi:10.1038/s41592-018-0235-4

47. 47. Finc K, Chojnowski M, Bonna K. fMRIDenoise: automated denoising, denoising strategies comparison, and functional connectivity data quality control. Published online 2019. https://zenodo.org/record/3383310#.Y5vBF3ZBy5c

48. Satterthwaite TD, Elliott MA, Gerraty RT, et al. An improved framework for confound regression and filtering for control of motion artifact in the preprocessing of resting-state functional connectivity data. NeuroImage. 2013;64:240–256. doi:10.1016/j.neuroimage.2012.08.052

49. Schaefer A, Kong R, Gordon EM, et al. Local-Global Parcellation of the Human Cerebral Cortex from Intrinsic Functional Connectivity MRI. Cerebral Cortex. 2018;28(9):3095–3114. doi:10.1093/cercor/bhx179

50. Oostenveld R, Fries P, Maris E, Schoffelen JM. FieldTrip: Open Source Software for Advanced Analysis of MEG, EEG, and Invasive Electrophysiological Data. Computational Intelligence and Neuroscience. 2011;2011:1–9. doi:10.1155/2011/156869

51. Thomas Yeo BT, Krienen FM, Sepulcre J, et al. The organization of the human cerebral cortex estimated by intrinsic functional connectivity. Journal of Neurophysiology. 2011;106(3):1125–1165. doi:10.1152/jn.00338.2011

52. Mesulam M. From sensation to cognition. Brain. 1998;121(6):1013–1052. doi:10.1093/brain/121.6.1013

53. tsne: t-Distributed Stochastic Neighbor Embedding. Published online 2022 2016. https://au.mathworks.com/help/stats/tsne.html#bvmb4f9

54. Kobak D, Linderman GC. Initialization is critical for preserving global data structure in both t-SNE and UMAP. Nat Biotechnol. 2021;39(2):156–157. doi:10.1038/s41587-020-00809-z

55. Nichols TE, Holmes AP. Nonparametric permutation tests for functional neuroimaging: A primer with examples. Hum Brain Mapp. 2001;15(1):1–25. doi:10.1002/hbm.1058

56. Oligschläger S, Huntenburg JM, Golchert J, Lauckner ME, Bonnen T, Margulies DS. Gradients of connectivity distance are anchored in primary cortex. Brain Struct Funct. 2017;222(5):2173–2182. doi:10.1007/s00429-016-1333-7

57. Wang Y, Royer J, Park B yong, et al. Long-Range Connections Mirror and Link Microarchitectural and Cognitive Hierarchies in the Human Brain. Neuroscience; 2021. doi:10.1101/2021.10.25.465692

58. Butler T, Weisholtz D, Isenberg N, et al. Neuroimaging of frontal-limbic dysfunction in schizophrenia and epilepsy-related psychosis: Toward a convergent neurobiology. Epilepsy Behav. 2012;23(2):113–122. doi:10.1016/j.yebeh.2011.11.004

59. Martínez K, Martínez-García M, Marcos-Vidal L, et al. Sensory-to-Cognitive Systems Integration Is Associated With Clinical Severity in Autism Spectrum Disorder. Journal of the American Academy of Child & Adolescent Psychiatry. 2020;59(3):422–433. doi:10.1016/j.jaac.2019.05.033

60. Lenka A, Jhunjhunwala KR, Saini J, Pal PK. Structural and functional neuroimaging in patients with Parkinson’s disease and visual hallucinations: A critical review. Parkinsonism & Related Disorders. 2015;21(7):683–691. doi:10.1016/j.parkreldis.2015.04.005

61. O’Callaghan C, Hall JM, Tomassini A, et al. Visual Hallucinations Are Characterized by Impaired Sensory Evidence Accumulation: Insights From Hierarchical Drift Diffusion Modeling in Parkinson’s Disease. Biological Psychiatry: Cognitive Neuroscience and Neuroimaging. 2017;2(8):680–688. doi:10.1016/j.bpsc.2017.04.007

62. Barnes J, Boubert L, Harris J, Lee A, David AS. Reality monitoring and visual hallucinations in Parkinson’s disease. Neuropsychologia. 2003;41(5):565–574. doi:10.1016/S0028-3932(02)00182-3

63. Zarkali A, Adams RA, Psarras S, Leyland LA, Rees G, Weil RS. Increased weighting on prior knowledge in Lewy body-associated visual hallucinations. Brain Communications. 2019;1(1):fcz007. doi:10.1093/braincomms/fcz007

64. Gallagher DA, Parkkinen L, O’Sullivan SS, et al. Testing an aetiological model of visual hallucinations in Parkinson’s disease. Brain. 2011;134(11):3299–3309. doi:10.1093/brain/awr225

65. Montagnese M, Vignando M, ffytche D, Mehta MA. Cognitive and visual processing performance in Parkinson’s disease patients with vs without visual hallucinations: A meta-analysis. Cortex. 2022;146:161–172. doi:10.1016/j.cortex.2021.11.001

66. Muller AJ, O’Callaghan C, Walton CC, Shine JM, Lewis SJG. Retrospective Neuropsychological Profile of Patients With Parkinson Disease Prior to Developing Visual Hallucinations. J Geriatr Psychiatry Neurol. 2017;30(2):90–95. doi:10.1177/0891988716686830

67. Miller EK, Cohen JD. An Integrative Theory of Prefrontal Cortex Function. Annu Rev Neurosci. 2001;24(1):167–202. doi:10.1146/annurev.neuro.24.1.167

68. Spreng RN, Stevens WD, Chamberlain JP, Gilmore AW, Schacter DL. Default network activity, coupled with the frontoparietal control network, supports goal-directed cognition. Neuroimage. 2010;53(1):303–317. doi:10.1016/j.neuroimage.2010.06.016

69. Bar M. Visual objects in context. Nat Rev Neurosci. 2004;5(8):617–629. doi:10.1038/nrn1476

70. Kveraga K, Boshyan J, Bar M. Magnocellular Projections as the Trigger of Top-Down Facilitation in Recognition. J Neurosci. 2007;27(48):13232–13240. doi:10.1523/JNEUROSCI.3481-07.2007

71. ffytche DH, Creese B, Politis M, et al. The psychosis spectrum in Parkinson disease. Nat Rev Neurol. 2017;13(2):81–95. doi:10.1038/nrneurol.2016.200

72. Chaudhuri KR, Schapira AH. Non-motor symptoms of Parkinson’s disease: dopaminergic pathophysiology and treatment. 2009;8:11.

73. Iarkov A, Barreto GE, Grizzell JA, Echeverria V. Strategies for the Treatment of Parkinson’s Disease: Beyond Dopamine. Front Aging Neurosci. 2020;12:4. doi:10.3389/fnagi.2020.00004

74. O’Brien J, Taylor JP, Ballard C, et al. Visual hallucinations in neurological and ophthalmological disease: pathophysiology and management. J Neurol Neurosurg Psychiatry. 2020;91(5):512–519. doi:10.1136/jnnp-2019-322702

75. Powell A, Ireland C, Lewis SJG. Visual Hallucinations and the Role of Medications in Parkinson’s Disease: Triggers, Pathophysiology, and Management. JNP. 2020;32(4):334–343. doi:10.1176/appi.neuropsych.19110316

76. Meltzer HY, Mills R, Revell S, et al. Pimavanserin, a Serotonin2A Receptor Inverse Agonist, for the Treatment of Parkinson’s Disease Psychosis. Neuropsychopharmacol. 2010;35(4):881–892. doi:10.1038/npp.2009.176

77. Girn M, Roseman L, Bernhardt B, Smallwood J, Carhart-Harris R, Spreng RN. Serotonergic psychedelic drugs LSD and psilocybin reduce the hierarchical differentiation of unimodal and transmodal cortex. NeuroImage. Published online April 2022:119220. doi:10.1016/j.neuroimage.2022.119220

78. Shine JM, O’Callaghan C, Walpola IC, et al. Understanding the effects of serotonin in the brain through its role in the gastrointestinal tract. Brain. 2022;145(9):2967–2981. doi:10.1093/brain/awac256

79. O’Callaghan C, Hezemans FH, Ye R, et al. Locus coeruleus integrity and the effect of atomoxetine on response inhibition in Parkinson’s disease. Brain. 2021;144(8):2513–2526. doi:10.1093/brain/awab142

