## Supplementary Material for "Abnormal higher-order network interactions in Parkinson’s disease visual hallucinations"

**1. fMRI preprocessing**	Page 2

**2. Rationale for using t-SNE**	Page 4

**3. t-SNE simulations and verification**	Page 5

**1. fMRI preprocessing**

**Anatomical data preprocessing**

The T1-weighted (T1w) image was corrected for intensity non-uniformity (INU) with N4BiasFieldCorrection (Tustison et al. 2010), distributed with ANTs 2.3.3 (Avants et al. 2008, RRID:SCR_004757), and used as T1w-reference throughout the workflow. The T1w-reference was then skull-stripped with a Nipype implementation of the antsBrainExtraction.sh workflow (from ANTs), using OASIS30ANTs as target template. Brain tissue segmentation of cerebrospinal fluid (CSF), white-matter (WM) and gray-matter (GM) was performed on the brain-extracted T1w using fast (FSL 5.0.9, RRID:SCR_002823, Zhang, Brady, and Smith 2001). Volume-based spatial normalization to two standard spaces (MNI152NLin2009cAsym, MNI152NLin6Asym) was performed through nonlinear registration with antsRegistration (ANTs 2.3.3), using brain-extracted versions of both T1w reference and the T1w template. The following templates were selected for spatial normalization: ICBM 152 Nonlinear Asymmetrical template version 2009c [Fonov et al. (2009), RRID:SCR_008796; TemplateFlow ID: MNI152NLin2009cAsym], FSL’s MNI ICBM 152 non-linear 6th Generation Asymmetric Average Brain Stereotaxic Registration Model [Evans et al. (2012), RRID:SCR_002823; TemplateFlow ID: MNI152NLin6Asym].

**Functional data preprocessing**

First, a reference volume and its skull-stripped version were generated using a custom methodology of fMRIPrep. Susceptibility distortion correction (SDC) was omitted. The BOLD reference was then co-registered to the T1w reference using flirt (FSL 5.0.9, Jenkinson and Smith 2001) with the boundary-based registration (Greve and Fischl 2009) cost-function. Co-registration was configured with nine degrees of freedom to account for distortions remaining in the BOLD reference. Head-motion parameters with respect to the BOLD reference (transformation matrices, and six corresponding rotation and translation parameters) are estimated before any spatiotemporal filtering using mcflirt (FSL 5.0.9, Jenkinson et al. 2002). The BOLD time-series (including slice-timing correction when applied) were resampled onto their original, native space by applying the transforms to correct for head-motion. These resampled BOLD time-series will be referred to as preprocessed BOLD in original space, or just preprocessed BOLD.

The BOLD time-series were resampled into standard space, generating a preprocessed BOLD run in MNI152NLin2009cAsym space. First, a reference volume and its skull-stripped version were generated using a custom methodology of fMRIPrep. Automatic removal of motion artifacts using independent component analysis (ICA-AROMA, Pruim et al. 2015) was performed on the preprocessed BOLD on MNI space time-series after removal of non-steady state volumes and spatial smoothing with an isotropic, Gaussian kernel of 6mm FWHM (full-width half-maximum). Corresponding “non-aggresively” denoised runs were produced after such smoothing. Additionally, the “aggressive” noise-regressors were collected and placed in the corresponding confounds file.

Several confounding time-series were calculated based on the preprocessed BOLD: framewise displacement (FD), DVARS and three region-wise global signals. FD was computed using two formulations following Power (absolute sum of relative motions, Power et al. (2014)) and Jenkinson (relative root mean square displacement between affines, Jenkinson et al. (2002)). FD and DVARS are calculated for each functional run, both using their implementations in Nipype (following the definitions by Power et al. 2014). The three global signals are extracted within the CSF, the WM, and the whole-brain masks. Additionally, a set of physiological regressors were extracted to allow for component-based noise correction (CompCor, Behzadi et al. 2007). Principal components are estimated after high-pass filtering the preprocessed BOLD time-series (using a discrete cosine filter with 128s cut-off) for the two CompCor variants: temporal (tCompCor) and anatomical (aCompCor). tCompCor components are then calculated from the top 2% variable voxels within the brain mask. For aCompCor, three probabilistic masks (CSF, WM and combined CSF+WM) are generated in anatomical space. The implementation differs from that of Behzadi et al. in that instead of eroding the masks by 2 pixels on BOLD space, the aCompCor masks are subtracted a mask of pixels that likely contain a volume fraction of GM. This mask is obtained by thresholding the corresponding partial volume map at 0.05, and it ensures components are not extracted from voxels containing a minimal fraction of GM.

Finally, these masks are resampled into BOLD space and binarized by thresholding at 0.99 (as in the original implementation). Components are also calculated separately within the WM and CSF masks. For each CompCor decomposition, the k components with the largest singular values are retained, such that the retained components’ time series are sufficient to explain 50 percent of variance across the nuisance mask (CSF, WM, combined, or temporal). The remaining components are dropped from consideration. The head-motion estimates calculated in the correction step were also placed within the corresponding confounds file. The confound time series derived from head motion estimates and global signals were expanded with the inclusion of temporal derivatives and quadratic terms for each (Satterthwaite et al. 2013). Frames that exceeded a threshold of 0.5 mm FD or 1.5 standardised DVARS were annotated as motion outliers. All resamplings can be performed with a single interpolation step by composing all the pertinent transformations (i.e., head-motion transform matrices, susceptibility distortion correction when available, and co-registrations to anatomical and output spaces). Gridded (volumetric) resamplings were performed using antsApplyTransforms (ANTs), configured with Lanczos interpolation to minimize the smoothing effects of other kernels (Lanczos 1964). Non-gridded (surface) resamplings were performed using mri_vol2surf (FreeSurfer).

Many internal operations of fMRIPrep use Nilearn 0.6.2 (Abraham et al. 2014, RRID:SCR_001362), mostly within the functional processing workflow. For more details of the pipeline, see [the section corresponding to workflows in fMRIPrep’s documentation](https://fmriprep.readthedocs.io/en/latest/workflows.html).

**2. Rationale for using t-SNE**

Functional connectivity is the correlation between fMRI timeseries for different regions of the brain. Regions that activate at similar timings will have higher correlations between them, and regions with dissimilar activity timing will have lower correlations. t-Stochastic Neighbour Embedding (t-SNE), can take functional connectivity as an input and calculate a simplified similarity score of functional connectivity in lower dimensions (2-3), prioritising relationships between regions in the original space. The following similarity score can be visualised in a coordinate space such that regions sharing similar connectivity profiles are placed closer together and regions with dissimilar connectivity profiles are more distant from each other. As each region can be described by a set of coordinates (x, y, z), we can calculate the Euclidean distance between regions as a proxy of functional connectivity in the t-SNE embedding space. Regions with similar connectivity profiles will have *smaller* distances between each other and regions with dissimilar connectivity profiles will have *greater* distances between each other.

In MATLAB, the t-SNE algorithm (tsne, Copyright 2016-2019 The MathWorks, Inc.) can be employed with the following parameters for adjustment:

***Algorithm (Barneshut, exact*):** Barnes-Hut approximation is an alternative algorithm for t-SNE developed to improve memory efficiency, making t-SNE calculations practical for large datasets. The alternative option (‘exact’), optimises the Kullback-Leibler divergence of distributions between the original and embedded space. The Kullback-Leibler divergence considers how different the distribution of one probability distribution is different from another when calculating the similarity scores.

***Distance:*** Specifies which distance metric is used to separate data points.

***Embedding dimensions:*** Number of dimensions for data visualisation (2 or 3).

***PCA Initialisation:*** Reducing the dimensions of the original data through Principal Component Analysis (PCA) and selecting the number of components to include in the t-SNE algorithm. PCA initialisation provides consistency to t-SNE output (Kobak and Linderman, 2021) and also increases computation efficiency by reducing the dimensions. Appropriate number of components was decided by running False Nearest Neighbours (FNN) and choosing the smallest number of components that resulted in <0.1% false neighbours.

***Perplexity:*** Approximates how many local neighbours should be assigned to each point. Changing perplexity affects how many surrounding points should be considered neighbouring to a datapoint. Setting large values of perplexity would encourage denser clusters, while smaller values would result in distributed sparse clusters.

***Learning Rate:*** The effect of learning on the optimisation process. Lower learning rates decrease the influence of learning during earlier optimisation processes. Higher learning rates retain learning even in later optimisation processes.

***Exaggeration:*** Cluster size. Changing exaggeration affects the size of the clusters as well as the space between clusters. The t-SNE algorithm uses exaggeration in the first 99 optimisation iterations to facilitate cluster formation (Belkina *et al.*, 2019).

**3. t-SNE Simulations and verification**

To find t-SNE plots that represent the functional connectivity data in a meaningful way, we ran the t-SNE algorithm on a 7T fMRI data of healthy controls (n = 59). The algorithm was run across 11 iterations of perplexity (1-100), learning rate (1-1000), and exaggeration (1-100) each for a total of 1331 iterations. As t-SNE randomly generates the starting coordinates, a specific seed was set for consistency and reproducible results. The Euclidean distance was calculated between each region (n = 400) and every other region resulting in a region by region (400 x 400) Euclidean distance map for each t-SNE plot. As mentioned previously, the Euclidean distance is a measurement of functional similarity between regions where regions that are different have a larger distance between them, and regions that are similar have smaller distances between them.

***Yeo Network Ratio:*** As a measurement of how well the t-SNE output represents established networks from resting-state fMRI, we created a ratio that compares connections within and between networks from the Yeo 7-network atlas (Yeo *et al.*, 2011). An average *within* network score was computed by averaging the Euclidean distance for edges connecting regions within the same network (i.e., visual region to visual region). An average *between* networks score was computed by averaging the Euclidean distance for edges connecting regions from different networks (i.e., visual region to somatomotor region). These scores were combined into a ratio by dividing the average within network score by the average between network score (Equation 1). Ratio scores greater than 1 occur when the Euclidean distance *within* networks is greater than the distance *between* networks, suggesting less distinct networks. For our analyses we preferred lower ratios that indicated more distinct networks.

$$Yeo Network Ratio= \frac{mean ED within networks}{mean ED between networks} (1)$$

ED = Euclidean distance

Networks = Yeo 7-network atlas

***Low- to High-dimension distance correlation:*** To check whether the t-SNE outputs were a faithful representation of the original embedding, we calculated the Pearson’s correlation between each region in the t-SNE distance map, with the corresponding region from the distance map in the original embedding. The distance map in the original embedding (400 x 400) was produced by calculating the Euclidean distance for each region to every other region. The Pearson’s correlation between the two maps was calculated for each region resulting in a correlation coefficient for each region (n = 400). The average correlation was used to summarise how well a t-SNE map represented the original embedding, where higher correlations indicated a more faithful representation.

**K-means clustering and adjusted mutual information (AMI)**: To identify the range of values for each parameter that produces an ‘ideal’ t-SNE representation of the data, we applied k-means clustering on the correlation scores. K-means clustering aims to create clusters while minimising within-cluster variances (Hartigan, 1975). K-means clustering was applied for a range of possible k clusters (2-20) with 100 iterations for each number of k clusters. To identify numbers of clusters that were consistent, the Adjusted Mutual Information (AMI) (Vinh and Epps, 2009; Vinh, Epps and Bailey, 2009, 2010) was calculated between iterations of k clusters. The average AMI for each k cluster was visualised. For the correlation scores, specifying k = 5 clusters resulted in the most consistent results. Each of the five clusters were visualised and interpreted and the cluster with the highest correlation score and the lowest Yeo Network Ratio was chosen as the ‘ideal’ t-SNE plot. From these results, the chosen cluster included t-SNE plots (n = 27) with correlation scores >= 0.83 and a Yeo Network Ratio score <= 0.54.

**Comparing ‘ideal’ t-SNE plots to other plots:** A quality check of whether the cluster chosen was different to the other clusters is to compare the similarity of the distance maps within the chosen cluster and between the chosen cluster and distance maps outside the cluster (Böhm, Berens and Kobak, 2021). From our chosen cluster, we randomly sampled five t-SNE plots and correlated their t-SNE maps between each other (5 per plot). We also randomly sampled five t-SNE plots from outside the chosen cluster and correlated each t-SNE plot from our chosen cluster with each of the plots outside of the cluster (5 per plot). The average correlations for each plot were visualised, and permutation testing was conducted comparing correlations within a cluster against correlations between clusters (5000 permutations). Indeed, the correlations within the cluster were significantly higher than the correlations with plots between clusters (p < 0.05).

**Final t-SNE plots**

From the parameter sweep and cluster analysis, the chosen cluster was dependent on a high perplexity (80-100) and was insensitive to changes in learning rate and exaggeration. For further t-SNE analysis conducted in the Parkinson’s disease dataset, the following values were set for each parameter:

- Algorithm = barneshut
- Distance = Euclidean
- NumDimensions = 3
- NumPCAComponents = 3
- Perplexity = 90
- LearnRate = 500
- Exaggeration = 50

**Limitations**

The t-SNE cluster identified from our analyses was specific to our question and the analysis pipeline we had in place. By no means is this cluster the ‘correct’ global representation of brain functional connectivity. We advise that before conducting t-SNE analyses in a dataset, one first follows our steps in designing a stringent criterion to test the t-SNE plots against before deciding on a set of parameter values to use. Furthermore, as the t-SNE algorithm does not project plots onto the exact same starting positions, in order to compare t-SNE plots directly we recommend aligning individual t-SNE plots to the population average (see procrustes, Copyright 1993-2009 The MathWorks, Inc.).

**References**

Abraham, Alexandre, *et al.* (2014) “Machine Learning for Neuroimaging with Scikit-Learn.” Frontiers in Neuroinformatics 8. <https://doi.org/10.3389/fninf.2014.00014>.

Avants, B.B., C.L. Epstein, M. Grossman, and J.C. Gee (2008) “Symmetric Diffeomorphic Image Registration with Cross-Correlation: Evaluating Automated Labeling of Elderly and Neurodegenerative Brain.” Medical Image Analysis 12 (1): 26–41. <https://doi.org/10.1016/j.media.2007.06.004>.

Behzadi, Yashar, Khaled Restom, Joy Liau, and Thomas T. Liu. (2007) “A Component Based Noise Correction Method (CompCor) for BOLD and Perfusion Based fMRI.” *NeuroImage* 37 (1): 90–101. <https://doi.org/10.1016/j.neuroimage.2007.04.042>.

Belkina, A.C. *et al.* (2019) ‘Automated optimized parameters for T-distributed stochastic neighbor embedding improve visualization and analysis of large datasets’, *Nature Communications*, 10(1), p. 5415. Available at: https://doi.org/10.1038/s41467-019-13055-y.

Böhm, J.N., Berens, P. and Kobak, D. (2021) ‘Attraction-Repulsion Spectrum in Neighbor Embeddings’. arXiv. Available at: http://arxiv.org/abs/2007.08902 (Accessed: 15 May 2022).

Esteban, Oscar, *et al.* (2018) “FMRIPrep.” *Software*. Zenodo. <https://doi.org/10.5281/zenodo.852659>.

Esteban, Oscar, *et al.* (2018) “fMRIPrep: A Robust Preprocessing Pipeline for Functional MRI.” *Nature Methods*. <https://doi.org/10.1038/s41592-018-0235-4>.

Evans, AC, *et al.* (2012) “Brain Templates and Atlases.” *NeuroImage* 62 (2): 911–22. <https://doi.org/10.1016/j.neuroimage.2012.01.024>.

Fonov, VS, *et al.* (2009) “Unbiased Nonlinear Average Age-Appropriate Brain Templates from Birth to Adulthood.” *NeuroImage* 47, Supplement 1: S102. <https://doi.org/10.1016/S1053-8119(09)70884-5>.

Gorgolewski, K., *et al.* (2011) “Nipype: A Flexible, Lightweight and Extensible Neuroimaging Data Processing Framework in Python.” *Frontiers in Neuroinformatics* 5: 13. <https://doi.org/10.3389/fninf.2011.00013>.

Gorgolewski, Krzysztof J. *et al*. (2018) “Nipype.” *Software*. Zenodo. <https://doi.org/10.5281/zenodo.596855>.

Greve, Douglas N, and Bruce Fischl (2009) “Accurate and Robust Brain Image Alignment Using Boundary-Based Registration.” *NeuroImage* 48 (1): 63–72. <https://doi.org/10.1016/j.neuroimage.2009.06.060>.

Hartigan, J.A. (1975) *John A. Hartigan - Clustering Algorithms-John Wiley & Sons (1975).pdf*. John Wiley & Sons.

Jenkinson, Mark, *et al.* (2002) “Improved Optimization for the Robust and Accurate Linear Registration and Motion Correction of Brain Images.” *NeuroImage* 17 (2): 825–41. <https://doi.org/10.1006/nimg.2002.1132>.

Jenkinson, Mark, and Stephen Smith (2001) “A Global Optimisation Method for Robust Affine Registration of Brain Images.” *Medical Image Analysis* 5 (2): 143–56. <https://doi.org/10.1016/S1361-8415(01)00036-6>.

Kobak, D. and Linderman, G.C. (2021) ‘Initialization is critical for preserving global data structure in both t-SNE and UMAP’, *Nature Biotechnology*, 39(2), pp. 156–157. Available at: https://doi.org/10.1038/s41587-020-00809-z.

Lanczos, C. (1964) “Evaluation of Noisy Data.” *Journal of the Society for Industrial and Applied Mathematics Series B Numerical Analysis* 1 (1): 76–85. <https://doi.org/10.1137/0701007>.

Power, Jonathan D. *et al.* (2014) “Methods to Detect, Characterize, and Remove Motion Artifact in Resting State fMRI.” *NeuroImage* 84 (Supplement C): 320–41. <https://doi.org/10.1016/j.neuroimage.2013.08.048>.

Pruim, Raimon H. R. *et al.* (2015) “ICA-AROMA: A Robust ICA-Based Strategy for Removing Motion Artifacts from fMRI Data.” *NeuroImage* 112 (Supplement C): 267–77. <https://doi.org/10.1016/j.neuroimage.2015.02.064>.

Satterthwaite, T.D. *et al.* (2013) ‘An improved framework for confound regression and filtering for control of motion artifact in the preprocessing of resting-state functional connectivity data’, *NeuroImage*, 64, pp. 240–256. Available at: https://doi.org/10.1016/j.neuroimage.2012.08.052.

Thomas Yeo, B.T. *et al.* (2011) ‘The organization of the human cerebral cortex estimated by intrinsic functional connectivity’, *Journal of Neurophysiology*, 106(3), pp. 1125–1165. Available at: https://doi.org/10.1152/jn.00338.2011.

Tustison, N. J. *et al.* (2010) “N4ITK: Improved N3 Bias Correction.” *IEEE Transactions on Medical Imaging* 29 (6): 1310–20. <https://doi.org/10.1109/TMI.2010.2046908>.

Vinh, N.X. and Epps, J. (2009) ‘A Novel Approach for Automatic Number of Clusters Detection in Microarray Data Based on Consensus Clustering’, in *2009 Ninth IEEE International Conference on Bioinformatics and BioEngineering*. *2009 Ninth IEEE International Conference on Bioinformatics and BioEngineering (BIBE)*, Taichung, Taiwan: IEEE, pp. 84–91. Available at: https://doi.org/10.1109/BIBE.2009.19.

Vinh, N.X., Epps, J. and Bailey, J. (2009) ‘Information Theoretic Measures for Clusterings Comparison: Is a Correction for Chance Necessary?’, p. 8.

Vinh, N.X., Epps, J. and Bailey, J. (2010) ‘Information Theoretic Measures for Clusterings Comparison: Variants, Properties, Normalization and Correction for Chance’, p. 18.

Zhang, Y., M. Brady, and S. Smith, (2001) “Segmentation of Brain MR Images Through a Hidden Markov Random Field Model and the Expectation-Maximization Algorithm.” *IEEE Transactions on Medical Imaging* 20 (1): 45–57. <https://doi.org/10.1109/42.906424>.
